# Evaluation of urine lipoarabinomannan and host response signatures for culture-negative tuberculosis using composite criteria and Bayesian Latent Class Analysis

**DOI:** 10.64898/2026.11.18.26368340

**Authors:** Robert Reiss, Kattya Lopez, William Honnen, Nisha Modi, Lesly Suarez Caceres, Nandini Dendukuri, Ian Schiller, Vaishnavi Kaipilyawar, Christie Eichberg, Lydia Nakiyingi, Chad Centner, Susan E. Dorman, Jerrold Ellner, W. Evan Johnson, Padmini Salgame, Abraham Pinter, Yingda L. Xie

## Abstract

**Background:** Culture-negative TB is a major diagnostic challenge, and the lack of a reliable reference standard hinders evaluation of non-sputum diagnostic tests. We compared composite reference criteria and Bayesian latent class analysis to examine how different culture-negative TB definitions affect disease classification and evaluations of two non-sputum biomarkers: urine lipoarabinomannan (uLAM) and host transcriptional response signatures (HRS).

**Methods:** Urine and blood samples from 112 sputum culture-negative and 55 culture-positive symptomatic adults evaluated for tuberculosis from South Africa, Uganda, and Kenya, alongside 17 controls with other respiratory conditions, were analyzed using uLAM and HRS assays. Sensitivity and specificity were estimated for each assay based on three reference criteria: composite microbiological, composite clinical, and a latent class model.

**Results:** Positive classification proportions were 0.29 (32/112) and 0.46 (51/112) for the composite criteria and 0.20 (22/112) for the latent class model, with the Bayesian model estimating a prevalence of 0.31 (95%CrI 0.11-0.64). Sensitivity and specificity for uLAM ranged from 0.06–0.13 and 0.92–0.96, respectively. Sensitivity and specificity for HRS ranged from 0.47–0.78 and 0.56–0.69, respectively. uLAM results were significantly associated with microbiological positivity (OR=5.38, 95%CI=1.63–13.02).

**Conclusion:** Three distinct reference frameworks identified different individuals as having culture-negative TB while producing index assay sensitivity and specificity estimates with overlapping confidence and credible intervals, leading to similar conclusions regarding biomarker utility. This supports the use of complementary reference approaches to assess robustness to disease classification uncertainty in heterogeneous conditions such as culture-negative TB.

**Summary:** We evaluated potential sputum culture-negative TB using complementary diagnostic criteria and Bayesian latent class models, revealing extensive molecular and clinical diversity. Similar performance of two non-sputum diagnostic assays across reference frameworks supports triangulation when no single diagnostic gold standard exists.

## INTRODUCTION

Approximately 2.4 million incident tuberculosis (TB) cases, caused by *Mycobacterium tuberculosis* (Mtb), are underreported or underdiagnosed yearly, implying substantial hidden disease burden and transmission.^1^ Conventional sputum culture-based diagnostics do not capture the full scope of active TB, particularly among individuals without microbiologic confirmation at presentation. At least 15-20% of newly diagnosed cases are sputum culture-negative TB (Cx⁻TB) with diagnosis based on clinical presentation, radiographic evidence, treatment response or non-culture molecular assay.^2,3^ Evidence from non-sputum-based assessments suggests that Cx⁻TB may constitute over 20% of undiagnosed symptomatic evaluees in some settings,^4,5^ may progress to microbiologically-confirmed disease,^6,7^ and may associate with viable Mtb in aerosols in over 80-90% of cases.^4,8^ Yet Cx⁻TB diagnostic development remains constrained by the lack of a reference standard.

Recently developed non-sputum-based diagnostics use urine or blood samples.^9,10^ Urine lipoarabinomannan (uLAM) lateral flow assays generated interest as point-of-care (POC) diagnostics with value among people living with HIV-1 with immunosuppression, but possessed limited sensitivity in HIV-1 negative and non-immunosuppressed populations.^11^ The WHO recommended these assays for the subset of HIV-positive people with advanced HIV disease or signs and symptoms of TB,^12,13^ based on results from the Determine TB LAM Ag assay, a rapid, POC lateral flow urine test produced by Alere (currently Abbott) that uses undefined polyclonal antibodies to capture and quantitate the antigen.^14^ Advanced versions of these tests are currently being evaluated for broader use.^15^

The identification of high affinity monoclonal antibodies (mAbs) against LAM has allowed improvements in the sensitivity and specificity of these assays.^16–18^ Newer assays utilize a pair of high affinity mAbs; S4-20, that recognizes an MTX-dependent epitope present in the capping domain of LAM from Mtb and other slow-growing mycobacteria,^19–22^ and A194-01, specific for a broadly conserved Ara4/Ara6-dependent epitope.^16,23^ These mAbs have also been incorporated into a new POC test, the Fujifilm SILVAMP TB LAM assay, that includes a proprietary silver amplification step that further improves assay sensitivity.^24–26^ However, despite their greater sensitivity, the ability of the newer immunoassays to detect TB LAM in urine from patients without HIV-1 coinfections was insufficient to meet the WHO’s requirements for a global screening assay. Recent results from the Pinter laboratory have shown that using the IgM isotype of A194-01 in place of the standard IgG improves sensitivity for uLAM detection.^23^

Host immune response signature (HRS) assays for Mtb infection have shown promise in identifying prevalent TB^27,28^ or paucibacillary disease^29^ and predicting progression,^30^ although with limited ability to distinguish TB from other respiratory conditions (ORC).^31^ Divergent performance profiles between uLAM and HRS assays reflect the detection of different disease signals – pathogen burden and host immune activation – and raises the possibility of joint use in multi-modal diagnostic strategies. However, development of such multimodal strategies is hindered by the lack of a Cx⁻TB diagnostic gold standard. Previous guidance has recommended evaluating nonsputum TB biomarkers against multiple reference standards and using latent class models to account for uncertainty in disease classification.^32^ but the consequences of applying this approach to Cx⁻TB remain poorly characterized.

We evaluate the diagnostic performance of uLAM and HRS assays for prevalent Cx⁻TB in symptomatic adults in Kenya, South Africa, and Uganda using three reference criteria. We compare results from each criterion and explore the use of composite reference standards and Bayesian mixture models to diagnose Cx⁻TB.

## METHODS

### Study populations

#### Enrollment, cohort definitions, and baseline and follow-up procedures

Symptomatic adults evaluated for pulmonary TB in Kenya, South Africa and Uganda (Ultra cohort) were consecutively enrolled as previously described.^33^ Baseline sputum, blood and urine were collected for sputum smear microscopy, Ultra and GeneXpert MTB/RIF (Xpert) testing, and Lowenstein-Jensen (LJ) or mycobacteria growth indicator tube (MGIT) culture. Baseline MGIT culture incubation was extended from 6 to 10 weeks for culture-negative participants. Supplemented MGIT cultures were performed at month 2 only among participants who had not initiated TB treatment, as previously described. Blood was collected in PAXgene Blood RNA Tubes (BD) for HRS analysis. Remaining sputum, blood and urine were stored at −80°C.

Untreated individuals whose enrollment culture results were negative with or without other negative diagnostic results and who participated in a longitudinal sub-study were identified as described previously as sputum culture-negative (Cx⁻).^5^ Sputum and blood were collected at months 2, 6 and 12. MGIT culture, smear microscopy and Ultra were conducted at each follow-up to assess progression. For untreated participants without a positive baseline culture by standard methods, baseline cultures were incubated for up to 10 weeks, and culture-filtrate–supplemented MGIT cultures were performed at month 2.

The target condition was probable Cx⁻TB at baseline, whereby microbiological and clinical findings obtained during follow-up were considered supportive evidence to ascertain baseline disease status. Cx⁻ participants were classified as either “probable culture-negative TB” (positive) or “unlikely culture-negative TB” (negative) via three sets of complementary criteria: (i) a microbiological composite reference (MRC), (ii) a clinical composite reference (CRC) serving as an extension of the MRC, and (iii) using a Bayesian latent class model (LCM). Participants were classified positive under the MRC based on detection of Mtb mRNA transcripts at baseline, positive results from baseline culture extended from 6 weeks to 10 weeks, positive results from MGIT at any follow-up, positive results from supplemented culture at month 2 follow-up or positive Ultra at month 2 follow-up with symptom progression. Participants were classified positive under the CRC if they were positive under the MRC or if they were clinically diagnosed with TB and improved with treatment.

#### TB-positive and negative cohorts

To establish baseline sensitivity, 20 of 180 Mtb culture-positive smear-positive (Cx⁺Sm⁺) parent study participants were randomly selected as positive controls, along with 35 baseline MGIT positive, Ultra trace-to-low positive and smear-negative (Cx⁺Sm⁻) “paucibacillary” participants.

For negative controls, participants from a low-prevalence local adult population were enrolled at University Hospital (UH) in Newark, NJ between April 2021 and April 2024 under approved Rutgers eIRB protocol Pro2020001138 after obtaining written informed consent. Of 26 enrolled, 16 were selected following TB rule-out and diagnosis with ORC, including COPD (6), asthma only (3), pneumonia only (2), and 1 case each of Kaposi sarcoma, interstitial lung disease, acute respiratory failure, hypoxic respiratory failure and pulmonary *Mycobacterium avium* infection. One participant from the Ultra cohort was considered ORC after an alternative Kaposi sarcoma diagnosis.

### Biomarker evaluation

#### Longitudinal culture and progression, Mtb mRNA

Participants were considered longitudinal culture-positive if they were positive for Mtb in baseline extended cultures, month 2 supplemented MGIT cultures, or any follow-up (months 2, 6, 12) standard MGIT cultures. Mtb mRNA was extracted and tested as previously described.^5^

#### Urine LAM immunoassay

White 96-well luminometry plates (Greiner600) were coated with rabbit anti-methylthioxylose monoclonal antibody (S4-20, FIND). All blocking and binding steps used 2% blotto (non-fat dry milk in PBS containing NaN_3_). Following blocking, urine samples were plated in triplicate; bound uLAM was detected with biotinylated human anti-Arabinose 4 monoclonal IgM (A194-01), followed by HRP-conjugated streptavidin (Jackson ImmunoResearch). Chemiluminescence was measured as Relative Light Units (RLUs) using Elisa Bright substrate (Advansta) on a SpectraMax Mini luminometer (Molecular Devices). The signal-to-noise ratio was the mean RLU of triplicate sample wells divided by the mean RLU of background (blotto-only) wells. Positive/negative cutoffs of 2.15 for the signal-to-noise ratio were derived independently via ROC-AUC analysis separating 10 confirmed TB participants from 9 controls without evidence of TB disease (Infectious Diseases Institute, Makerere University, Kampala, Uganda), as well as from 9 ORC participants (Figure S1a).

Urine was additionally analyzed with the Fujifilm SILVAMP TB LAM (FujiLAM) assay as a comparator according to manufacturer instructions and results were rated by two or three blinded raters.

#### Host response signatures

Total RNA was extracted from blood using the PAXgene Blood RNA kit (Qiagen, Hilden, Germany). An 81-gene NanoString panel (NanoString Technologies) was performed as previously described.^34^ Non-parametric, unsupervised gene set variation analysis (GSVA) was used to generate signature scores.^35^ Diagnostic performance was evaluated with the R package TBSignatureProfiler.^36,37^

Positive/negative cutoffs were derived via ROC-AUC analysis separating 20 anonymously enrolled, US-based, TB-negative healthy controls from 30 sputum culture-positive participants randomly selected from the Ultra cohort (Figure S1b and S1c). The PREDICT29^37^ and ACS-COR 16^38^ signatures were chosen to determine HRS positivity due to their strong ability to separate healthy controls from sputum-positive participants.^5^ Participants with GSVA scores exceeding 0.5813 for ACS-COR 16 or less than –0.1048 for PREDICT29 were considered positive.

## Statistical analysis

### Statistical analysis

Statistical analyses were performed using R, version 4.5.0. Demographic and clinical statistics were calculated by cohort, TB status, and uLAM and HRS biomarker positivity (Tables 1, 2 and S1). Differences were analyzed via Kruskal-Wallis with post-hoc Dunn tests (Benjamini-Hochberg adjusted), Mann-Whitney U, Fisher’s exact test (Holm-Bonferroni adjusted) and McNemar’s test.

**Table 1.** Participant clinical characteristics for each cohort, combined and by culture and AFB smear status.

|  | **All participants<br>(n = 184)** | **Cx<sup>−</sup><br>(n = 112)** | **Cx<sup>+</sup>Sm<sup>+</sup><br>(n = 20)** | **Cx<sup>+</sup>Sm<sup>−</sup><br>(n = 35)** | **ORC<sup>1</sup><br>(n = 17)** | **p-<br>value\*** |
|---|---|---|---|---|---|---|
| Demographic characteristics |  |  |  |  |  |  |
| Sex (male) (no., %) | 107/184 (58.2) | 66/112 (58.9) | 13/20 (65.0) | 20/35 (57.1) | 8/17 (47.1) | 0.74 |
| Age (median, IQR) | 38 (31.0 – 48.0) | 38.5 (32.0 – 47.3) | 32.5 (27.0 – 42.0) | 35.0 (30.5 – 44.5) | 57 (43.0 – 61.0) | **0.001** |
| Study site (no., %) |  |  |  |  |  |  |
| Kenya | 75/184 (40.7) | 44/112 (39.3) | 10/20 (50.0) | 20/35 (57.1) | 1/17 (5.9) | **< 0.001** |
| South Africa | 56/184 (30.4) | 38/112 (33.9) | 6/20 (30.0) | 12/35 (34.3) | 0/17 (0.0) |  |
| Uganda | 37/184 (20.1) | 30/112 (26.8) | 4/20 (20.0) | 3/35 (8.6) | 0/17 (0.0) |  |
| United States | 16/184 (8.7) | 0/112 (0.0) | 0/20 (0.0) | 0/35 (0.0) | 16/17 (94.1) |  |
| Clinical characteristics |  |  |  |  |  |  |
| Prior history of TB (no., %) | 56/184 (30.4) | 41/112 (36.6) | 5/20 (25.0) | 5/35 (14.3) | 5/17 (29.4) | 0.08 |
| HIV positive status (no., %) | 104/184 (56.5) | 63/112 (56.3) | 11/20 (55.0) | 23/35 (65.7) | 7/17 (41.2) | 0.42 |
| TB symptom severity, baseline (no., %)<sup>1</sup> |  |  |  |  |  |  |
| Not ill | 35/168 (20.8) | 30/112 (26.8) | 3/20 (15.0) | 2/35 (5.7) | 0/1 (0.0) | **0.025** |
| Mildly ill | 93/168 (55.4) | 60/112 (53.6) | 8/20 (40.0) | 25/35 (71.4) | 0/1 (0.0) |  |
| Moderately ill | 32/168 (19.0) | 18/112 (16.1) | 7/20 (35.0) | 6/35 (17.1) | 1/1 (100.0) |  |
| Gravely ill | 8/168 (4.8) | 4/112 (3.6) | 2/20 (10.0) | 2/35 (5.7) | 0/1 (0.0) |  |
| Symptoms improved with treatment (no., %)<sup>2</sup> | 66/74 (89.2) | 30/36 (83.3) | 19/19 (100.0) | 16/18 (88.9) | 1/1 (100.0) | 0.23 |
| Microbiologic tests |  |  |  |  |  |  |
| AFB smear positive (no., %)<sup>1</sup> | 21/170 (12.4) | 1/112 (0.9) | 20/20 (100.0) | 0/35 (0.0) | 1/3 (33.3) | **< 0.001** |
| Progression to culture positive TB (no., %)<sup>1</sup> | 2/168 (1.2) | 2/112 (1.8) | 0/20 (0.0) | 0/35 (0.0) | 0/1 (0.0) | 1.00 |
| Extended MGIT culture positive (no., %)<sup>1,3</sup> | 0/102 (0.0) | 0/101 (0.0) | N/A | N/A | 0/1 (0.0) | N/A |
| Month 2 supplemented MGIT positive, (no., %)<sup>1,3</sup> | 5/69 (7.2) | 4/68 (5.9) | N/A | N/A | 1/1 (100.0) | 0.07 |
| Ultra status, baseline (no., %)<sup>1</sup> |  |  |  |  |  |  |
| Negative | 40/168 (23.8) | 40/112 (35.7) | 0/20 (0.0) | 0/35 (0.0) | 0/1 (0.0) | **< 0.001** |
| Trace | 53/168 (31.5) | 49/112 (43.8) | 0/20 (0.0) | 3/35 (8.6) | 1/1 (100.0) |  |
| Very Low | 26/168 (15.5) | 14/112 (12.5) | 1/20 (5.0) | 11/35 (31.4) | 0/1 (0.0) |  |
| Low | 33/168 (19.6) | 9/112 (8.0) | 3/20 (15.0) | 21/35 (60.0) | 0/1 (0.0) |  |
| Medium | 2/168 (1.2) | 0/112 (0.0) | 2/20 (10.0) | 0/35 (0.0) | 0/1 (0.0) |  |
| High | 14/168 (8.3) | 0/112 (0.0) | 14/20 (70.0) | 0/35 (0.0) | 0/1 (0.0) |  |
| mRNA transcript detected (no., %)<sup>1,4</sup> | 44/139 (31.7) | 25/112 (22.3) | 13/15 (86.7) | 6/11 (54.5) | 0/1 (0.0) | **< 0.001** |
| FujiLAM assay positive (no., %) | 24/184 (13.0) | 2/112 (1.8) | 13/20 (65.0) | 9/35 (25.7) | 0/17 (0.0) | **< 0.001** |
| Index tests |  |  |  |  |  |  |
| uLAM positive (no., %) | 24/184 (13.0) | 7/112 (6.3) | 9/20 (45.0) | 6/35 (17.1) | 2/17 (11.8) | **< 0.001** |
| HRS positive (no., %) | 93/184 (50.5) | 50/112 (44.6) | 18/20 (90.0) | 18/35 (51.4) | 7/17 (41.2) | **0.001** |

\*p-value is for the comparison versus participants that are negative for both biomarkers by Fisher’s exact test or Kruskal-Wallis test.
<sup>1</sup>Excludes 16 US-based ORC participants, who were not evaluated for symptom severity, tested by Ultra, tested for mRNA transcripts, or had longitudinal culture performed. Additionally, 14 were not tested by AFB smear.
<sup>2</sup>Among treated participants only; 110 participants were not started on treatment at any point.
<sup>3</sup>Among the Cx<sup>−</sup> cohort, extended MGIT cultures are missing for 11 and supplemented MGIT culture are missing for 44. Longitudinal culture was not performed for participants who had positive baseline culture.
<sup>4</sup>mRNA transcript results are missing for 5 Cx<sup>+</sup>Sm<sup>+</sup> and 24 Cx<sup>+</sup>Sm<sup>−</sup> participants.

**Table 2.** Baseline demographic and clinical characteristics for cohort Cx^−^, by diagnostic criterion.

|  | **Total** | **Microbiological Criterion** |  |  | **Clinical Criterion** |  |  | **LCM Classification** |  |  |
|---|---|---|---|---|---|---|---|---|---|---|
|  | **Cx–<br>(n = 112)** | **TB-positive<br>(n = 32)** | **TB-negative<br>(n = 80)** | **p-value\*** | **TB-positive<br>(n = 51)** | **TB-negative<br>(n = 61)** | **p-value\*** | **TB-positive<br>(n = 22)** | **TB-negative<br>(n = 90)** | **p-value\*** |
| Demographic characteristics |  |  |  |  |  |  |  |  |  |  |
| Sex (male) (no., %) | 66/112 (58.9) | 19/32 (59.4) | 47/80 (58.8) | 1.00 | 31/51 (60.8) | 35/61 (57.4) | 0.85 | 14/22 (63.6) | 52/90 (57.8) | 0.81 |
| Age (median, IQR) | 38.5 (32.0 – 47.3) | 39 (33.0 – 50.3) | 38 (29.8 – 45.0) | 0.18 | 40 (33.5 – 49.5) | 37 (29.0 – 45.0) | 0.14 | 40 (33.0 – 50.3) | 38.0 (32.0 – 45.0) | 0.40 |
| Study site (no., %) |  |  |  |  |  |  |  |  |  |  |
| Kenya | 44/112 (39.3) | 13/32 (40.6) | 31/80 (38.8) | 0.452 | 14/51 (27.5) | 30/61 (49.2) | **0.001** | 10/22 (45.5) | 34/90 (37.8) | 0.63 |
| South Africa | 38/112 (33.9) | 13/32 (40.6) | 25/80 (31.3) |  | 15/51 (29.4) | 23/61 (37.7) |  | 8/22 (36.4) | 30/90 (33.3) |  |
| Uganda | 30/112 (26.8) | 6/32 (18.8) | 24/80 (30.0) |  | 22/51 (43.1) | 8/61 (13.1) |  | 4/22 (18.2) | 26/90 (28.9) |  |
| United States | 0/112 (0.0) | N/A | N/A | N/A | N/A | N/A | N/A | N/A | N/A | N/A |
| Clinical characteristics |  |  |  |  |  |  |  |  |  |  |
| Prior history of TB (no., %) | 41/112 (36.6) | 17/32 (53.1) | 24/80 (30.0) | **0.03** | 26/51 (51.0) | 15/61 (24.6) | **0.006** | 11/22 (50.0) | 30/90 (33.3) | 0.22 |
| HIV positive status (no., %) | 63/112 (56.3) | 18/32 (56.3) | 45/80 (56.3) | 1.00 | 30/51 (58.8) | 33/61 (54.1) | 0.70 | 13/22 (59.1) | 50/90 (55.5) | 0.81 |
| TB symptom severity, baseline (no., %) |  |  |  |  |  |  |  |  |  |  |
| Not ill | 30/112 (26.8) | 5/32 (15.6) | 25/80 (31.3) | 0.16 | 16/51 (31.4) | 14/61 (23.0) | 0.16 | 3/22 (13.6) | 27/90 (30.0) | **0.030** |
| Mildly ill | 60/112 (53.6) | 20/32 (62.5) | 40/80 (50.0) |  | 28/51 (54.9) | 32/61 (52.5) |  | 12/22 (54.5) | 48/90 (53.3) |  |
| Moderately ill | 18/112 (16.1) | 5/32 (15.6) | 13/80 (16.3) |  | 5/51 (9.8) | 13/61 (21.3) |  | 3/22 (13.6) | 15/90 (16.7) |  |
| Gravely ill | 4/112 (3.6) | 2/32 (6.3) | 2/80 (2.5) |  | 2/51 (3.9) | 2/61 (3.3) |  | 4/22 (18.2) | 0/90 (0.0) |  |
| Symptoms improved with treatment (no., %)<sup>1</sup> | 30/36 (83.3) | 6/9 (66.6%) | 24/27 (88.9) | 0.15 | 25/28 (89.3) | 5/8 (62.5) | 0.11 | 4/6 (66.7) | 26/30 (86.7) | 0.26 |
| Microbiologic tests |  |  |  |  |  |  |  |  |  |  |
| AFB smear positive (no., %) | 1/112 (0.9) | 0/32 (0.0) | 1/80 (1.3) | 1.00 | 1/51 (2.0) | 0/61 (0.0) | 0.46 | 1/22 (4.5) | 0/90 (0.0) | 0.20 |
| Progression to culture positive TB (no., %) | 2/112 (1.8) | 2/32 (6.3) | 0/80 (0.0) | 0.08 | 2/51 (3.9) | 0/61 (0.0) | 0.21 | 2/22 (9.1) | 0/90 (0.0) | **0.037** |
| Extended MGIT culture positive (no., %)<sup>1</sup> | 0/101 (0.0) | 0/29 (0.0) | 0/72 (0.0) | 1.00 | 0/45 (0.0) | 0/56 (0.0) | 1.00 | 0/20 (0.0) | 0/81 (0.0) | 1.00 |
| Month 2 supplemented MGIT positive, (no., %)<sup>2</sup> | 4/68 (5.9) | 4/20 (20.0) | 0/48 (0.0) | **0.006** | 4/21 (19.0) | 0/47 (0.0) | **0.007** | 4/16 (25.0) | 0/52 (0.0) | **0.002** |
| Ultra status, baseline (no., %) |  |  |  |  |  |  |  |  |  |  |
| Negative | 40/112 (35.7) | 15/32 (46.9) | 25/80 (31.3) | 0.59 | 15/51 (29.4) | 25/61 (41.0) | **0.02** | 7/22 (31.8) | 33/90 (36.7) | 0.55 |
| Trace | 49/112 (43.8) | 8/32 (25.0) | 41/80 (51.3) |  | 19/51 (37.3) | 30/61 (49.2) |  | 9/22 (40.9) | 40/90 (44.4) |  |
| Very Low | 14/112 (12.5) | 6/32 (18.8) | 8/80 (10.0) |  | 11/51 (21.6) | 3/61 (4.9) |  | 5/22 (22.7) | 9/90 (10.0) |  |
| Low | 9/112 (8.0) | 3/32 (9.4) | 6/80 (7.5) |  | 6/51 (11.8) | 3/61 (4.9) |  | 1/22 (4.5) | 8/90 (8.9) |  |
| Medium | 0/112 (0.0) | 0/32 (0.0) | 0/80 (0.0) |  | 0/51 (0.0) | 0/61 (0.0) |  | 0/22 (0.0) | 0/90 (0.0) |  |
| High | 0/112 (0.0) | 0/32 (0.0) | 0/80 (0.0) |  | 0/51 (0.0) | 0/61 (0.0) |  | 0/22 (0.0) | 0/90 (0.0) |  |
| mRNA transcript detected (no., %) | 25/112 (22.3) | 25/32 (78.1) | 0/80 (0.0) | **< 0.001** | 25/51 (49.0) | 0/61 (0.0) | **< 0.001** | 0/22 (0.0) | 25/90 (27.8) | **0.003** |
| FujiLAM assay positive (no., %) | 2/112 (1.8) | 1/32 (3.1) | 1/80 (1.3) | 0.49 | 2/51 (3.9) | 0/61 (0.0) | 0.21 | 1/22 (4.5) | 1/90 (1.1) | 0.36 |
| Index tests |  |  |  |  |  |  |  |  |  |  |
| uLAM positive (no., %) | 7/112 (6.3) | 4/32 (12.5) | 3/80 (3.8) | 0.10 | 4/51 (7.8) | 3/61 (4.9) | 0.70 | 1/22 (4.5) | 6/90 (6.7) | 1.00 |
| HRS positive (no., %) | 50/112 (44.6) | 15/32 (46.9) | 35/80 (43.8) | 0.83 | 25/51 (49.0) | 25/61 (41.0) | 0.45 | 22/22 (100.0) | 28/90 (31.1) | **< 0.001** |

\*TB-positive vs TB-negative by Fisher’s exact test or Mann-Whitney U test.
<sup>1</sup>Among n = 36 patients that started on treatment during the study period.
<sup>2</sup>Among n = 102 patients with completed extended MGIT cultures and n = 69 patients with completed supplemented MGIT cultures.

Indicator sensitivities and specificities with Wilson 95% confidence intervals were calculated with respect to the composite reference standards, and prevalence and indicator accuracy metrics with 95% credible intervals (95%CrI) were estimated via LCM. Sensitivities and specificities were re-estimated using Cx⁺Sm⁺ and Cx⁺Sm⁻ samples as cases and ORC and healthy volunteer samples as controls. ROC curves were generated for independent cutoff derivation and re-derived sensitivities and specificities (Figure S2).

Counts and sensitivity and specificity estimates for the uLAM assay, by cohort and by reference standard, were further stratified by HIV status and CD4^+^ T-cell count (Tables S2 and S3).

Exploratory descriptive logistic regression was used to estimate the association of uLAM and HRS assays with TB status across a mixed cohort controlling for clinical covariates. Models were trained on 70% of participants (n=128) with TB status as the outcome and evaluated on a 30% test set (n=56). Explanatory variables included uLAM signal-to-noise ratio and HRS composite score (summed PREDICT29 and ACS-COR z-scores). Covariates included HIV status, prior TB history, symptom duration and total number of symptoms.

#### Bayesian LCM

A two-class Bayesian LCM was constructed assuming the two underlying latent classes are Cx⁻TB positive and no TB, using Ultra, mRNA qRT-PCR, longitudinal culture, uLAM and HRS as indicators (Figure S3). Model parameters include participant class, TB disease prevalence and assay sensitivity and specificity. The LCM was programmed in Just Another Gibbs Sampler (JAGS) and executed with *rjags*. Model optimization, troubleshooting and interpretation were performed with assistance from the Google Gemini large language model version 3 Pro.

Ultra, mRNA, longitudinal culture and uLAM sensitivities were assumed dependent on Mtb burden after conditioning on TB positive status. This was modeled using a shared individual-level random effect. Because HRS are driven by non-specific systemic inflammation rather than directly by Mtb burden, the shared Mtb burden effect was replaced with a separate random effect to accommodate individual-level variation in inflammation. Conditional independence was assumed in the TB-negative class. Missing data were imputed by JAGS assuming missing-at-random.

Informative prior distributions were chosen using previously estimated assay metrics among TB-positive Cx⁻ individuals (Table S4). Longitudinal culture specificity was treated as a constant of 98% based on culture specificity estimates.^39,40^

Markov Chain Monte Carlo Gibbs sampling was used to generate posterior distributions (three chains, 50,000 iterations). Parameters were estimated using posterior medians and 95% Credible Intervals (95%CrI), with individuals assigned to the Cx-TB positive class if their posterior probability of disease exceeded 50%. Sensitivity analysis assessed index test influence by comparing the full model to versions excluding index tests, including uLAM only, and including HRS only. After observing low sensitivity in the Mtb mRNA assay, the full model was re-run with priors corresponding to a range of possible mRNA sensitivities to determine how initial assumptions affected model outputs (Table S4).

## RESULTS

### Sample demographic and clinical characteristics

A total of 184 participants followed over 12 months as previously described^5^ were selected, including 112 sputum Cx⁻ (72 Ultra⁺ and 40 Ultra⁻) and 55 sputum culture-positive (Figure 1). Sample demographic and clinical statistics were calculated by cohort (Table 1). Of 184 participants, 107 (58.2%) were male and median age was 38. One ORC cohort participant diagnosed with pulmonary *Mycobacterium avium* infection returned a positive AFB smear. From the Cx^−^ cohort, extended MGIT results were missing for 11 (9.8%) participants and supplemented MGIT results were missing for 44 participants (39.3%) (Table S5). Missing extended MGIT results occurred alongside baseline culture contamination in 5 of 7 participants, whereas missing supplemented MGIT results reflected treatment initiation before protocol-specified month-2 testing in 26 participants.

**Figure 1.**
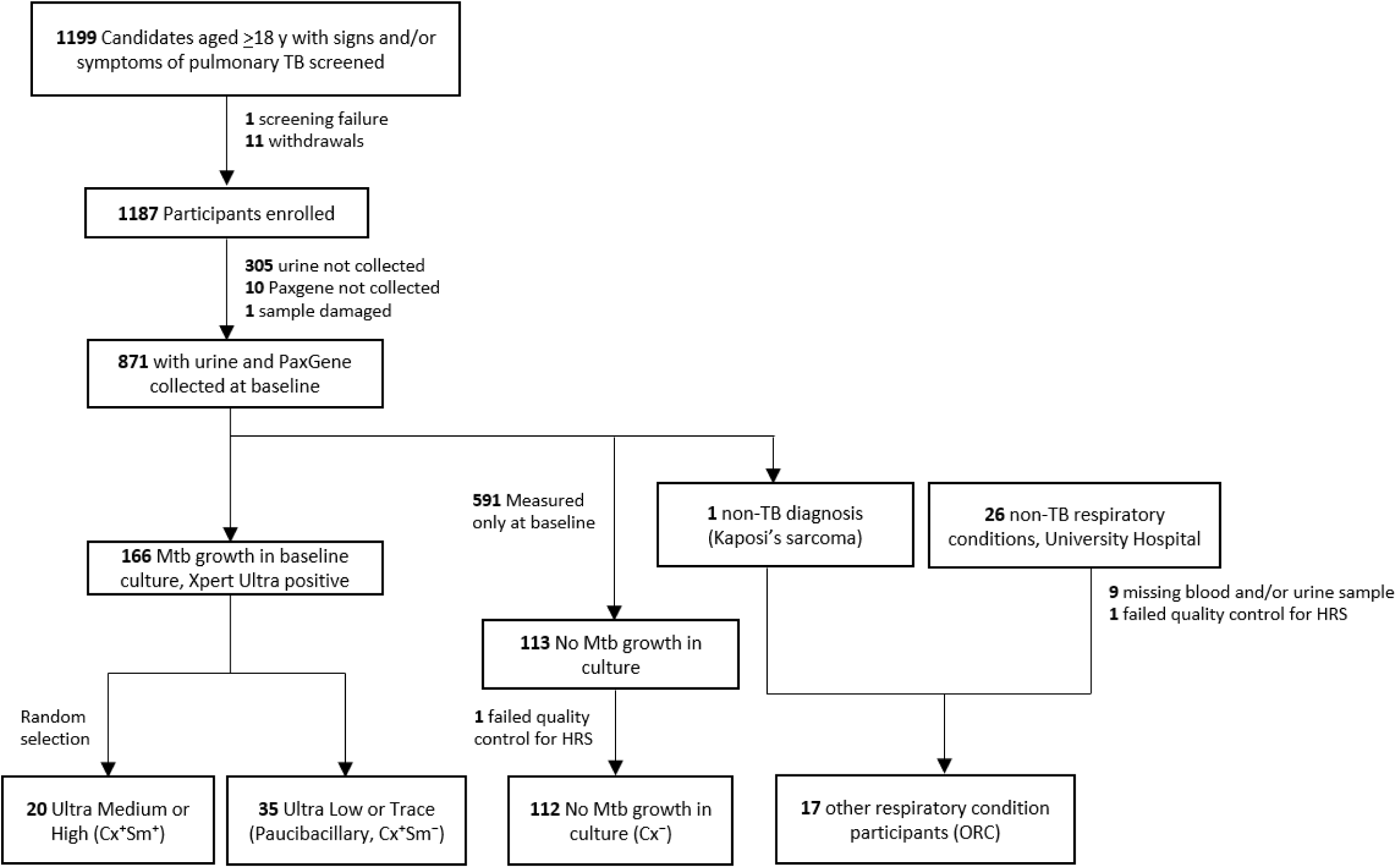
Study flowchart (STARD) of selection of participants for the cohort of interest, positive controls, healthy volunteers and ORC negative controls. Alt text: A STARD flowchart depicting study participants who were screened (n = 1199), enrolled (n = 1187), had samples collected (n = 871) and were assigned to Cx⁺Sm⁺ (n = 20), Cx⁻Sm⁻ (n = 35), Cx⁻ (n = 112) or ORC (n = 17) cohorts.

Statistics were calculated for the Cx^−^ cohort using each Cx⁻TB criterion (Table 2). Across all three criteria 20-46% of Cx⁻ participants were classified as TB-positive. The LCM estimated TB prevalence at 0.31 (95%CrI=0.11–0.64) and classified 22 of 112 participants (19.6%) as TB-positive. The proportion of participants considered “likely TB” under the CRC (51/112, 45.5%) is significantly higher than under both the MRC (32/112, 28.6%, McNemar p<0.001) and the LCM (McNemar p<0.001). MRC and LCM classification concordance is 64% (72/112) (Table S6).

Positive participants as defined by the MRC and CRC were significantly more likely to have prior history of TB and detectable mRNA in their sputum. Those classified positive by the CRC were significantly more likely to have higher semi-quantitative Ultra results compared to those classified as negative (p=0.02). Positive participants as defined by the LCM were significantly sicker and more likely to progress to active TB than those classified as negative.

### Diagnostic performance of uLAM and HRS index tests

uLAM assay estimates ranged from 0.06–0.13 sensitivity and 0.92–0.96 specificity (Table 3). No differences in uLAM sensitivity were observed by HIV status or immunocompromised status (Table S3). FujiLAM assay estimates ranged from 0.03–0.05 sensitivity and 0.99–1.00 specificity. HRS assay estimates ranged from 0.47–0.78 sensitivity and 0.56–0.69 specificity. Exploratory logistic regression analysis was used to evaluate index test associations with TB status in Cx⁻ participants (Table S7). In the model trained using the MRC to determine participant classification, uLAM results were significantly associated with TB status, with odds of positivity increasing by 5.38 for each one-unit increase in signal-to-noise ratio (OR=5.38, 95%CI=1.63–13.02, p=0.002).

**Table 3.** Estimates of sensitivity and specificity for the uLAM and HRS assays by contingency table analysis for microbiologic criterion, clinical criterion, and for the Bayesian LCM, with 95% Wilson confidence/credible intervals.

|  | **Microbiological criterion** |  |  | **Clinical criterion** |  |  | **Latent class model<sup>1</sup>** |  |
|---|---|---|---|---|---|---|---|---|
|  | **Sensitivity<br>(95% CI)** | **Specificity<br>(95% CI)** | **AUC<br>(95% CI<sup>2</sup>)** | **Sensitivity<br>(95% CI)** | **Specificity<br>(95% CI)** | **AUC<br>(95% CI<sup>2</sup>)** | **Sensitivity<br>(95% CrI)** | **Specificity<br>(95% CrI)** |
| **uLAM** | 0.13 (0.05–0.28) | 0.96 (0.90–0.99) | 0.66 (0.54–0.77) | 0.08 (0.03–0.18) | 0.95 (0.87–0.98) | 0.54 (0.43–0.65) | 0.06 (0.00–0.28) | 0.92 (0.82–0.98) |
| **FujiLAM** | 0.03 (0.01–0.16) | 0.99 (0.93–1.00) | – | 0.03 (0.01–0.13) | 1.00 (0.94–1.00) | – | – | – |
| **HRS** | 0.47 (0.31–0.64) | 0.56 (0.45–0.67) | – | 0.49 (0.36 – 0.62) | 0.59 (0.46–0.70) | – | 0.78 (0.34–0.99) | 0.69 (0.48–0.93) |
| **PREDICT29** | – | – | 0.53 (0.43–0.64) | – | – | 0.57 (0.46–0.67) | – | – |
| **ACS-COR** | – | – | 0.53 (0.41–0.64) | – | – | 0.51 (0.40–0.62) | – | – |
| **uLAM or HRS** | 0.56 (0.39–0.72) | 0.55 (0.44-0.65) | – | 0.55 (0.41–0.68) | 0.57 (0.45–0.69) | – | – | – |
| **uLAM and HRS** | 0.03 (0.01 – 0.16) | 0.98 (0.91 – 0.99) | – | 0.02 (0.003 – 0.10) | 0.97 (0.89 – 0.99) | – | – | – |

<sup>1</sup>Sensitivity and specificity parameters are estimated directly from the latent class model.
<sup>2</sup>From 2000 stratified bootstrap replicates.

### Sensitivity analysis

Sensitivity analysis evaluating LCM parameter stability with and without index tests showed confidence/credible interval overlap in prevalence, sensitivity and specificity estimates across the models (Table S8). Testing the influence of prior selection on mRNA assay sensitivity yielded estimates of 0.05 (95%CrI=0.00–0.24) for beta(0.77,4.23), 0.13 (95%CrI=0.02–0.42) for beta(1.85,3.15), 0.22 (95%CrI=0.06–0.76) for beta(2.93,2.07) and 0.63 (95%CrI=0.13–0.99) for beta(4.02, 0.98) (Table S9).

## DISCUSSION

To capture information on Cx⁻TB disease indicators without clear diagnostic criteria, we triangulated biomarker performance across two composite benchmarks and a Bayesian LCM. Sensitivity and specificity for individual assays were poor-to-modest (Table 3) across criteria. The largest estimate difference was a 0.31 HRS sensitivity increase between the MRC and LCM, though the parameter’s 95%CrI contains the MRC estimate. Despite differences in whom these three diagnostic frameworks classified as having culture-negative TB, estimates of biomarker performance were generally similar but imprecise due to high uncertainty. As such these frameworks should be treated as complementary approaches that do not definitively establish diagnostic accuracy.

Overall, the performance profiles of uLAM and HRS reflect the distinct biological signals captured by each assay. The uLAM assay’s very low sensitivity (0.06–0.13) and high specificity (0.92–0.96) reflect assay dependence on urinary antigen shedding as a proxy for bacillary burden — a signal absent in culture-negative disease outside of advanced immunosuppression, and that FujiLAM does not overcome. Another possible factor contributing to the lower sensitivity of the uLAM assay than that reported in previous studies is that sensitivity of the standard ELISA platform used in this study is limited by the background signal due to non-specific binding of antibodies to the polystyrene wells. This gives lower signal-to-noise ratios than that seen in the Meso-Scale platform, which is commonly used for these assays.^17,18,41–43^ In the Meso-Scale platform the target antigen is attached to carbon ink electrodes, which have higher surface areas than polystyrene wells and give inherently low backgrounds, and the electrical stimulation of light emission used in this system results in an amplified signal.

HRS showed higher sensitivity (0.47–0.78) but modest specificity (0.56–0.69) consistent with capturing systemic immune activation that persists with paucibacillary infection but failing to reliably discriminate TB from other inflammatory states. That both strategies underperformed reflects the nature of the population itself: Cx⁻TB here primarily associates with mild pulmonary disease with less illness severity – rather than disseminated TB – where antigen shedding and systemic immune activation may be minimal, placing these patients at the lower end of the detectable signal spectrum for both biomarker domains.

When combined using an "or" rule — reasoning that uLAM and HRS represent different and potentially complementary domains of TB biology — the combination yielded minimal improvement in sensitivity (0.56) over HRS alone while sacrificing specificity (0.55). Exploratory logistic regression corroborated the limited independent contribution of HRS: uLAM (OR=5.38, p=0.002) and symptom count (OR=1.74, p=0.002) were the only significant predictors under the MRC, with HRS contributing less discriminatory information than clinical symptoms alone. The combined model showed modest test-set performance (OR=3.74, p=0.03; sensitivity 0.54, specificity 0.77), driven by uLAM and symptom burden, and was non-significant against the CRC (OR=1.49, p=0.74).

Low concordance between the LCM and the CRC (0.54) and MRC (0.64) likely stems from bias inherent to composite criteria indicator selection.^44^ Because a perfect reference does not exist, requiring any positive test result for diagnosis increases sensitivity but decreases specificity, whereas requiring multiple positive tests increases specificity at the cost of sensitivity. Thus, different reasonable reference standards can identify substantially different groups of patients. The LCM avoids a fixed diagnostic rule by estimating disease status probabilistically from the observed data, although individual classifications still depend on the available indicators (Table S8). These findings suggest that a major obstacle to biomarker development for culture-negative TB is not simply finding a more sensitive test, but defining what constitutes disease when conventional microbiological confirmation is absent.

### Limitations

Due to the absence of a gold standard test, culture-negative samples were evaluated using different criteria, a strategy that cannot cover every possible diagnostic criterion. We also did not collect radiographic imaging as in studies that characterized Cx⁻TB.^45^ Because the uLAM and HRS assays are under development, cutoffs are not firmly established and vary depending on the samples selected for validation. Here, independently derived cutoffs were estimated with fewer paucibacillary samples, potentially inflating optimal cutoff estimations. Consequently, the selected cutoff may be overly conservative towards type I errors, especially for uLAM.

Despite similarity in LCM parameter estimates over varied initial assumptions, the selection of a two-class model produced outputs that may not fully align with established TB biology. Notably, positive mRNA results were absent among LCM-positive participants, reflecting an observed inverse association with longitudinal culture positivity in this cohort. Because longitudinal culture specificity was fixed at 98%, given the improbability of false positives from culture, the model mathematically associates negative mRNA results with membership in the TB-positive class. In a binary framework, the model correspondingly associates positive mRNA results with TB-negative status, outweighing positive results from the remaining indicators with lower or uninformative specificity priors. Nevertheless, both participants who progressed to culture-positive TB were classified as LCM-positive, providing microbiological support for the model.

While sensitivity analysis yielded a narrow range of prevalence estimates (0.31–0.37) across differing assumptions, individual class assignment showed much more volatility, ranging from 5% to 38% positive classification. This instability is explained by a high proportion of borderline assignments, where 37 participants (33%) had posterior probabilities of TB-positive class membership between 0.4 and 0.6. Changes in the model structure result in small shifts in posterior probability that cross the 0.5 decision threshold. These findings underscore the uncertainty in individual LCM class assignment for this cohort under varying model specifications, and for this reason the model should be considered complementary to the other two reference criteria.

Missing data in the extended and supplemented MGIT cultures also contributes to uncertainty in the classifications. Because treatment decisions influenced supplemented-culture availability, missingness may have been associated with underlying TB status, potentially affecting microbiological classification and latent class estimates.

### Conclusion

Evaluating assay performance for Cx⁻TB is difficult given the lack of standardized diagnostic criteria. Practically, Cx⁻TB has no observable ground truth, and different assays capture different slices of a variable disease state. Those identified by uLAM were consistent with a high mycobacterial burden, whereas HRS detected a broader, less specific inflammatory phenotype. We observed 15 unique biomarker patterns in the Cx⁻ cohort (Figure S4 and S5), suggesting extensive molecular and clinical diversity. We found that a triangulated framework combining biologically interpretable criteria with stress-tested LCMs provides one approach to evaluating Cx⁻TB without systematically privileging a single biomarker class. These findings support evaluating candidate biomarkers against multiple complementary definitions of Cx⁻TB to determine whether conclusions are robust to uncertainty in disease classification.

## Acknowledgements

We would like to thank the following collaborators for their contribution to this project; Alok Choudhary and Afsal Kolloli for serving as raters for the FujiLAM assay.

The Google Gemini large language model version 3 Pro was used in the optimization, troubleshooting and interpretation of the latent class model.

## Data Availability Statement

All quantitative data generated and analyzed in the course of this study and photographs of the FujiLAM assay results will be provided upon reasonable request.

## Author Contributions

RR: statistical data analysis and interpretation, LCM model development, data visualization and manuscript writing

KL: processed PAXgene blood samples and performed the HRS analysis

WH: performed the uLAM analysis

NM: processed sputum samples and conducted Mtb mRNA transcriptional analysis

LSC: enrolled ORC participants and managed sample storage and organization

VK: contributed to the HRS analysis and prepared data

ND: provided support for the LCM analysis

IS: provided support for the LCM analysis

CE: clinical data acquisition and interpretation

LN: clinical data acquisition and interpretation

CC: microbiological testing, clinical data acquisition and interpretation

SED: data acquisition, analysis and interpretation

JE: study design, analysis and interpretation

WEJ: developed the HRS machine learning pipeline

PS: study design, supervising HRS analysis and interpretation

AP: study design, developed the uLAM test, supervising the uLAM testing and interpretation, contributed to the writing the manuscript

YLX: conceptualization, study design, supervising analysis and overall interpretation All authors were involved in reviewing, editing and finalizing the manuscript.

## Figures, Tables, Data and Supplementary Materials

### Main Body Figures and Tables

**Figure 1.** Study flowchart (STARD) of selection of participants for the cohort of interest, positive controls, healthy volunteers and ORC negative controls

**Table 1.** Participant demographic and clinical characteristics for each cohort, by culture status and AFB smear status.

**Table 2.** Cohort 2 demographic and clinical characteristics, TB-positive vs. TB-negative, for microbiological criterion, clinical criterion, and latent class model classification (n = 112).

**Table 3.** Sensitivity and specificity estimates for index tests by the microbiological criterion, clinical criterion and latent class model classification.

