## Supplemental Materials for "Evaluation of urine lipoarabinomannan and host response signatures for culture-negative tuberculosis using composite criteria and Bayesian Latent Class Analysis"

Table of Contents

| Item | Contents | Page |
|---|---|---|
| *Latent Class Model Materials* | | |
| Item 1 | Latent class model specification | 1 |
| Item 2 | JAGS code for the latent class model. | 3 |
| Item 3 | Parameter estimates, quantiles, effective sample sizes and Gelman-Rubin statistics. | 6 |
| *Supplemental Tables* | | |
| Table S1 | Participant demographic characteristics, stratified by uLAM and HRS biomarker positivity. | 7 |
| Table S2 | uLAM and HRS positivity stratified by cohort, by HIV status and $CD4^+$ T-cell count. | 9 |
| Table S3 | uLAM and HRS positivity stratified by reference standard, by HIV status and $CD4^+$ T-cell count. | 11 |
| Table S4 | Prior distribution selection for latent class model parameters. | 14 |
| Table S5 | Missing data summary for indicator tests for the culture-negative cohort. | 16 |
| Table S6 | Concordance between clinical criterion, microbiological criterion and latent class classifications | 17 |
| Table S7 | Logistic regression model results | 18 |
| Table S8 | Sensitivity and specificity estimates for all indicator tests for four latent class models. | 19 |
| Table S9 | Class assignment and prevalence, sensitivity and specificity estimates of index tests under four assumed mRNA assay sensitivity priors. | 20 |
| *Supplemental Figures* | | |
| Figure S1 | ROC curves for uLAM and HRS assays (independently derived) | 21 |
| Figure S2 | ROC curves for uLAM and HRS assays (for culture positive samples and ORC samples) | 22 |
| Figure S3 | Latent class analysis DAG. | 23 |
| Figure S4 | Venn diagrams of index biomarker overlap by cohort and criterion. | 24 |
| Figure S5 | Venn diagram and upset plots of biomarker overlap for culture-negative TB cohort. | 25 |
| Figure S6 | Host response signature violin plots for all other signatures | 27 |
| Figure S7 | Host response signature violin plots for PREDICT29 and ACS-COR | 28 |
| Figure S8 | uLAM signal-to-noise ratio box plot. | 29 |
| Figure S9 | Latent class model MCMC posterior density plots | 30 |
| Figure S10 | Expanded STARD diagram. | 31 |

1. Latent class model specification.

Let $y_{ij} \in \{0, 1\}$ represent the observed binary test result for participant $i$ $(i = 1, 2 \ldots 112)$ on diagnostic test $j$ $(j = 1, 2 \ldots 5)$. Let $LC_i \in \{1, 2\}$ denote the unobserved latent class membership for participant $i$ (class 1 = TB-positive, class 2 = TB-negative).

Class membership is defined hierarchically via the prevalence parameter $\pi$:

$$\pi \sim Beta(2, 3)$$

$$LC1_i \sim Bernoulli(\pi)$$

$$LC_i \sim 2 - LC1_i$$

Conditional on class membership $LC_i$, test results $y_{ij}$ are Bernoulli distributed as follows:

$$y_{ij} \sim \text{Bernoulli}(p_{LC_i,i,j})$$

The conditional probabilities of a positive observation for each diagnostic test are modeled on the probit scale:

$$\text{Latent class 1:} \qquad p_{1,i,j} = \Phi(a_{1,j} + b.RE_j \cdot r_i)$$

$$\text{Latent class 2:} \qquad p_{2,i,j} = \Phi(a_{2,j})$$

where conditional dependence is modeled via the addition of random effects. Individual level random effects are normally distributed as $r_i \sim N(0, 1)$, and the test-level random effect parameter $b.RE$ is shared between tests 1, 2, 3 and 4, and independently estimated for test 5 as follows:

$$b.RE_1 = b.RE_2 = b.RE_3 = b.RE_4 \sim N(0, 1)|(b.RE_1 > 0)$$

$$b.RE_5 \sim N(0, 1)|(b.RE_5 > 0)$$

The probit intercepts $a_{1,j}$ and $a_{2,j}$ are parameterized in terms of test sensitivities and specificities as follows:

$$\text{Latent class 1:} \qquad a_{1,j} = probit(se_j) \cdot \sqrt{1 + b.RE_j^2} \text{ for } j = 1, 2, \ldots, 5$$

$$\text{Latent class 2:} \qquad a_{2,j} = -probit(sp_j) \text{ for } j \neq 3 \text{ and } a_{2,3} = -probit(0.98)$$

Prior distributions are specified as follows:

$$se_1 \sim Beta(13.90, 16.32)$$

$$se_2 \sim Beta(1.85, 3.15)$$

$$se_3 \sim Beta(8.94, 42.01)$$

$$se_4, se_5, sp_4, sp_5 \sim Beta(1, 1)$$

$$sp_1 \sim Beta(3.41, 0.59)$$

$$sp_2 \sim Beta(7.12, 1.33)$$

$sp_3 = 0.98$ (fixed)

2. JAGS code for the full latent class model.

```
# Outcomes of interest are the sensitivities and specificities for HRS
and LAM tests
# and prediction of TB status for each subject in the culture negative
(Cx-) cohort

### variable definitions ###

    # Variables
      # LC[i]: latent class membership/Mtb status LC for subject i (1 =
positive, 2 = negative)
      #    pLC: true underlying class probability
      #          pLC[1]: probability of true positive
      #    L: number of latent classes (L = 2, defined in R script)
      # N: number of participants (Cx- cohort, n = 112, defined in R
script)
      # i: ith subject 1-112
      # j: jth test 1-5
      #    J: number of tests (J = 5, defined in R script)
      # y: test result and covariate matrix
      # p: proportion of results (probability of positive test result
for subject i and test j)
      # se: sensitivity with regard to measurand
      # sp: specificity with regard to measurand

###----------------------###

model {
  for(i in 1:N) {

 ##############################
 ## Likelihood specification ##
 ##############################

      for(j in 1:J) {
           y[i,j] ~ dbin(p[LC[i],i,j], 1)
     }

      LC1[i] ~ dbern(pLC[1])
      LC[i] <- 2 - LC1[i]

      r[i] ~ dnorm(0,1)

##------------------------------------------------------------##
    ## Conditional probability of +ve observation for given latent
class ##
##------------------------------------------------------------------
##

      ## Latent class 1 - Mtb infection positive (Mtb+)
        p[1,i,1] <- phi(a[1,1] + b.RE[1] * r[i])
```

```
        p[1,i,2] <- phi(a[1,2] + b.RE[2] * r[i])
        p[1,i,3] <- phi(a[1,3] + b.RE[3] * r[i])
        p[1,i,4] <- phi(a[1,4] + b.RE[4] * r[i])
        p[1,i,5] <- phi(a[1,5] + b.RE[5] * r[i])

      ## Latent class 2 - Mtb infection negative (Mtb-)
        p[2,i,1] <- phi(a[2,1])
        p[2,i,2] <- phi(a[2,2])
        p[2,i,3] <- phi(a[2,3])
        p[2,i,4] <- phi(a[2,4])
        p[2,i,5] <- phi(a[2,5])
      }

#########################
## Prior distributions ##
#########################

  ##-----------------------------##
  ## Random effects parameters ##
  ##-----------------------------##

      a[1,1] <- probit(se[1])*sqrt(1 + b.RE[1]^2)
      a[1,2] <- probit(se[2])*sqrt(1 + b.RE[2]^2)
      a[1,3] <- probit(se[3])*sqrt(1 + b.RE[3]^2)
      a[1,4] <- probit(se[4])*sqrt(1 + b.RE[4]^2)
      a[1,5] <- probit(se[5])*sqrt(1 + b.RE[5]^2)

      a[2,1] <- -probit(sp[1])
      a[2,2] <- -probit(sp[2])
      a[2,3] <- -probit(0.98)
      a[2,4] <- -probit(sp[4])
      a[2,5] <- -probit(sp[5])

      b.RE[1] ~ dnorm(0,1)T(0,)
      b.RE[2] <- b.RE[1]
      b.RE[3] <- b.RE[1]
      b.RE[4] <- b.RE[1]
      b.RE[5] ~ dnorm(0,1)T(0,)

  ##------------------------------##
  ## Latent class probabilities ##
  ##------------------------------##

      pLC[1] ~ dbeta(2,3)

  ##-------------------------##
  ## Parameters of interest ##
  ##-------------------------##

     ## Sensitivity and specificity with regards to TB disease status

      #Sensitivity
```

```
            se[1] ~ dbeta(13.90, 16.32)
            se[2] ~ dbeta(1.85, 3.15)
            se[3] ~ dbeta(8.94, 42.01)
            se[4] ~ dbeta(1,1)
            se[5] ~ dbeta(1,1)

        #Specificity
            sp[1] ~ dbeta(3.41, 0.59)
            sp[2] ~ dbeta(7.12, 1.33)
            sp[4] ~ dbeta(1,1)
            sp[5] ~ dbeta(1,1)

      ## Prevalences ##

        #Prevalence of active TB disease
            prev <- pLC[1]

  }
```

3. Parameter estimates, quantiles, effective sample sizes and Gelman-Rubin statistics.

| | | | Quantiles | | | | |
|---|---|---|---|---|---|---|---|
| **Parameter** | **Code** | **Mean (SD)** | **2.5th** | **50th** | **97.5th** | **Eff. Sample Size** | **GR Statistic** |
| $Cx^{-}$TB prevalence | prev | 0.33 (0.14) | 0.11 | 0.31 | 0.64 | 3622 | 1.001 |
| Ultra sensitivity | se[1] | 0.45 (0.10) | 0.30 | 0.45 | 0.60 | 34045 | 1.000 |
| mRNA sensitivity | se[2] | 0.15 (0.10) | 0.02 | 0.13 | 0.42 | 5831 | 1.002 |
| Longitudinal culture | se[3] | 0.17 (0.05) | 0.09 | 0.17 | 0.28 | 12972 | 1.000 |
| uLAM sensitivity | se[4] | 0.08 (0.08) | 0.00 | 0.06 | 0.28 | 5296 | 1.001 |
| HRS sensitivity | se[5] | 0.75 (0.18) | 0.34 | 0.78 | 0.99 | 4039 | 1.002 |
| Ultra specificity | sp[1] | 0.65 (0.08) | 0.51 | 0.65 | 0.81 | 13623 | 1.002 |
| mRNA specificity | sp[2] | 0.73 (0.07) | 0.58 | 0.74 | 0.87 | 8727 | 1.001 |
| uLAM specificity | sp[4] | 0.92 (0.04) | 0.82 | 0.92 | 0.98 | 10386 | 1.005 |
| HRS specificity | sp[5] | 0.70 (0.11) | 0.48 | 0.69 | 0.93 | 5414 | 1.001 |
| Random effect (bacillary burden) | b.RE[1] | 0.38 (0.31) | 0.01 | 0.31 | 1.16 | 10944 | 1.001 |
| Random effect (host inflammation) | b.RE[5] | 0.84 (0.61) | 0.04 | 0.73 | 2.27 | 17233 | 1.000 |

### SUPPLEMENTAL FIGURES AND TABLES

**Table S1.** Baseline demographic and clinical characteristics by uLAM and HRS biomarker positivity.

| | **All participants (n = 184)** | **Negative for both biomarkers (n = 85)** | **uLAM positive (n = 24)** | **p-value*** | **HRS positive (n = 93)** | **p-value*** | **uLAM or HRS positive (n = 99)** | **p-value*** | **uLAM and HRS positive (n = 18)** | **p-value*** |
|---|---|---|---|---|---|---|---|---|---|---|
| Demographic characteristics | | | | | | | | | | |
| Sex (male) (no., %) | 107/184 (58.2) | 50/85 (58.8) | 12/24 (50.0) | 0.49 | 54/93 (58.1) | 1.00 | 57/99 (57.6) | 0.88 | 9/18 (50.0) | 0.60 |
| Age (median, IQR) | 38 (31.0 – 48.0) | 38 (31.0 – 46.0) | 35 (29.8 – 42.0) | 0.21 | 38 (31.0 – 48.0) | 0.94 | 39 (31.5 – 48.0) | 0.78 | 33 (27.5 – 37.3) | **0.01** |
| Study site (no., %) | | | | | | | | | | |
| Kenya | 75/184 (40.7) | 34/85 (40.0) | 12/24 (50.0) | 0.60 | 39/93 (41.9) | 0.054 | 41/99 (41.4) | 0.09 | 10/18 (55.6) | 0.23 |
| South Africa | 56/184 (30.4) | 20/85 (23.5) | 7/24 (29.2) | | 35/93 (37.6) | | 36/99 (36.4) | | 6/18 (33.3) | |
| Uganda | 37/184 (20.1) | 23/85 (27.1) | 4/24 (16.7) | | 12/93 (12.9) | | 14/99 (14.1) | | 2/18 (11.1) | |
| United States | 16/184 (8.7) | 8/85 (9.4) | 1/24 (4.2) | | 7/93 (7.5) | | 8/99 (8.1) | | 0/18 (0.0) | |
| Clinical characteristics | | | | | | | | | | |
| Prior history of TB (no., %) | 56/184 (30.4) | 23/85 (27.1) | 7/24 (29.2) | 0.80 | 30/93 (32.3) | 0.51 | 33/93 (35.5) | 0.42 | 4/18 (22.2) | 0.78 |
| HIV positive status (no., %) | 104/184 (56.4) | 40/85 (47.1) | 17/24 (70.8) | 0.06 | 61/93 (65.6) | **0.02** | 64/99 (64.6) | **0.02** | 14/18 (77.8) | **0.02** |
| TB symptom severity, baseline (no., %)[a] | | | | | | | | | | |
| Not ill | 35/168 (20.8) | 24/77 (31.2) | 4/23 (17.4) | 0.47 | 9/86 (10.5) | 1.00 | 11/91 (12.1) | 0.89 | 2/18 (11.1) | 0.31 |
| Mildly ill | 93/168 (55.4) | 44/77 (57.1) | 10/23 (43.5) | | 47/86 (54.7) | | 49/91 (53.8) | | 8/18 (44.4) | |
| Moderately ill | 32/168 (19.0) | 9/77 (11.7) | 6/23 (26.1) | | 22/86 (25.6) | | 23/91 (25.3) | | 5/18 (27.8) | |
| Gravely ill | 8/168 (4.8) | 0/77 (0.0) | 3/23 (13.0) | | 8/86 (9.3) | | 8/91 (8.8) | | 3/18 (16.7) | |
| Symptoms improved with treatment (no., %)[b] | 66/74 (89.2) | 26/32 (81.3) | 14/14 (100.0) | 0.16 | 38/40 (95.0) | 0.13 | 40/42 (95.2) | 0.07 | 12/12 (100.0) | 0.17 |
| Microbiologic tests | | | | | | | | | | |
| AFB smear positive (no., %)[a] | 21/170 (12.4) | 3/79 (3.8) | 9/23 (39.1) | **< 0.001** | 19/86 (22.1) | **< 0.001** | 19/91 (20.9) | **< 0.001** | 9/18 (50.0) | **< 0.001** |
| Progression to culture positive TB[a] (no., %) | 2/168 (1.2) | 0/77 (0.0) | 0/23 (0.0) | 1.00 | 2/86 (2.3) | 0.50 | 2/91 (2.2) | 0.50 | 0/18 (0.0) | 1.00 |
| Extended MGIT culture positive (no., %)[a,c] | 0/102 (0.0) | 0/51 (0.0) | 0/7 (0.0) | 1.00 | 0/46 (0.0) | 1.00 | 0/51 (0.0) | 1.00 | 0/2 (0.0) | 1.00 |
| Month 2 supplemented MGIT positive, (no., %)[a,c] | 5/69 (7.2) | 0/34 (0.0) | 1/6 (16.7) | 0.15 | 4/31 (12.9) | **0.046** | 5/35 (14.3) | 0.054 | 0/2 (0.0) | 1.00 |
| Ultra status, baseline (no., %)[a] | | | | | | | | | | |
| Negative | 40/168 (23.8) | 20/77 (26.0) | 5/23 (21.7) | 0.26 | 17/86 (19.8) | 0.63 | 20/91 (22.0) | 0.57 | 2/18 (11.1) | 0.17 |
| Trace | 53/168 (31.5) | 30/77 (39.0) | 3/23 (13.0) | | 21/86 (24.4) | | 23/91 (25.3) | | 1/18 (5.6) | |

| | | | | | | | | | | |
|---|---|---|---|---|---|---|---|---|---|---|
| Very Low | 26/168 (15.5) | 12/77 (15.6) | 1/23 (4.3) | | 14/86 (16.3) | | 14/91 (15.4) | | 1/18 (5.6) | |
| Low | 33/168 (19.6) | 13/77 (16.9) | 6/23 (26.1) | | 20/86 (23.3) | | 20/91 (22.0) | | 6/18 (33.3) | |
| Medium | 2/168 (1.2) | 0/77 (0.0) | 2/23 (8.7) | | 2/86 (2.3) | | 2/91 (2.2) | | 2/18 (11.1) | |
| High | 14/168 (8.3) | 2/77 (2.6) | 6/23 (26.1) | | 12/86 (14.0) | | 12/91 (13.2) | | 6/18 (33.3) | |
| mRNA transcript detected (no., %)[a,d] | 44/139 (31.7) | 17/62 (27.4) | 12/18 (66.7) | **0.004** | 24/72 (33.3) | 0.57 | 27/77 (35.1) | 0.36 | 9/13 (69.2) | **0.01** |
| FujiLAM assay positive (no., %) | 24/184 (13.0) | 2/85 (2.4) | 13/24 (54.2) | **< 0.001** | 22/93 (23.7) | **< 0.001** | 22/99 (22.2) | **< 0.001** | 13/18 (72.2) | **< 0.001** |

*p-value is for the comparison versus participants that are negative for both biomarkers by Fisher's exact test or Mann-Whitney U test.

[a]Among 16 US-based ORC participants, none were evaluated for symptom severity, tested by Ultra, tested for mRNA transcripts, or had longitudinal culture performed. Additionally, 14 were not tested by AFB smear.

[b]Among treated participants only; 110 participants were not started on treatment at any point.

[c]Among the $Cx^-$ cohort, extended MGIT cultures are missing for 11 and supplemented MGIT culture are missing for 44. Longitudinal culture was not performed for participants who had positive baseline culture.

[d]mRNA transcript results are missing for five $Cx^+Sm^+$ and 24 $Cx^+Sm^-$ participant

**Table S2.** Estimates of percent of positive uLAM and HRS assay results for each cohort, stratified by HIV status and $CD4^+$ T-cell count, with 95% confidence intervals.

| | **All participants (n = 184)** | **$Cx^-$ (n = 112)** | **$Cx^+Sm^+$ (%) (n = 20)** | **$Cx^+Sm^-$ (n = 35)** | **ORC[a] (n = 17)** | **p-value*** |
|---|---|---|---|---|---|---|
| **uLAM positive (no., %)** | 24/184 (13.0) [8.7 – 19.0] | 7/112 (6.3) [2.8 – 12.9] | 9/20 (45.0) [23.8 – 68.0] | 6/35 (17.1) [7.2 – 34.3] | 2/17 (11.8) [2.1 – 37.7] | **< 0.001** |
| **uLAM by HIV status** | | | | | | |
| HIV-positive (no., %) | 17/104 (16.3) [10.1 – 25.2] | 4/63 (6.3) [2.1 – 16.3] | 6/11 (54.5) [24.6 – 81.9] | 5/23 (21.7) [8.3 – 44.2] | 2/7 (28.6) [5.1 – 69.7] | **< 0.001** |
| HIV-negative (no., %) | 7/80 (8.8) [3.9 – 17.7] | 3/49 (6.1) [1.6 – 17.9] | 3/9 (33.3) [9.0 – 69.1] | 1/12 (8.3) [0.4 – 40.2] | 0/10 (0.0) [0.0 – 34.4] | 0.06 |
| **uLAM by $CD4^+$ T-cell count[b]** | | | | | | |
| < 200 (no., %) | 10/45 (22.2) [11.7 – 37.5] | 1/20 (5.0) [0.2 – 26.9] | 4/6 (66.7) [24.1 – 94.0] | 4/15 (26.7) [8.9 – 55.2] | 1/4 (25.0) [1.3 – 78.1] | **0.011** |
| ≥ 200 unit (no., %) | 6/58 (10.3) [4.3 – 21.8] | 3/43 (7.0) [1.8 – 20.1] | 2/5 (40.0) [7.3 – 83.0] | 1/8 (12.5) [0.7 – 53.3] | 0/2 (0.0) [0.0 – 80.2] | 0.13 |
| **HRS positive (no., %)** | 93/184 (50.5) [43.1 – 57.9] | 50/112 (44.6) [35.3 – 54.3] | 18/20 (90.0) [66.9 – 98.2] | 18/35 (51.4) [34.3 – 68.3] | 7/17 (41.2) [19.4 – 66.5] | **0.001** |
| **HRS by HIV status** | | | | | | |
| HIV-positive (no., %) | 61/104 (58.7) [48.6 – 68.1] | 34/63 (54.0) [41.0 – 66.4] | 11/11 (100.0) [67.9 – 1.00] | 14/25 (56.0) [35.3 – 75.0] | 2/7 (28.6) [5.1 – 69.7] | **0.006** |
| HIV-negative (no., %) | 32/80 (40.0) [29.4 – 51.6] | 16/49 (32.7) [20.4 – 47.7] | 7/9 (77.8) [40.2 – 96.1] | 4/12 (33.3) [11.3 – 64.6] | 5/10 (50.0) [23.7 – 76.3] | 0.07 |
| **HRS by $CD4^+$ T-cell count[b]** | | | | | | |
| < 200 (no., %) | 35/45 (77.8) [62.5 – 88.3] | 17/20 (85.0) [61.1 – 96.0] | 6/6 (100.0) [51.7 – 1.00] | 11/15 (73.3) [44.8 – 91.1] | 1/4 (25.0) [1.3 – 78.1] | **0.047** |
| ≥ 200 unit (no., %) | 26/58 (44.8) [32.0 – 58.4] | 17/43 (39.5) [25.4 – 55.5] | 5/5 (100.0) [46.4 – 1.00] | 3/8 (37.5) [10.2 – 74.1] | 1/2 (50.0) [9.5 – 90.5] | 0.06 |

$Cx^-$: sputum culture negative. $Cx^+Sm^+$: sputum culture positive, smear positive. $Cx^+Sm^-$: sputum culture positive, smear negative. ORC: other respiratory condition. uLAM: urine LAM assay. HRS: host response signature assay (n = 17)*Comparison across all the $Cx^{-}$, $Cx^{+}Sm^{+}$, $Cx^{+}Sm^{-}$ and ORC cohorts by Fisher’s exact test.

[a]One ORC participant is missing a $CD4^{+}$ measurement.

[b]Among HIV-positive participants only.

**Table S3.** Estimates of percent of positive uLAM and HRS assay results for Cx⁻ participants, stratified by HIV status and CD4⁺ T-cell count, with 95% confidence intervals.

| | Microbiological criterion | | Clinical criterion | | LCA Classification | |
|---|---|---|---|---|---|---|
| | **MRS+ (n = 32)** | **MRS- (n = 80)** | **CRS+ (n = 51)** | **CRS- (n = 61)** | **LCM+ (n = 22)** | **LCM- (n = 90)** |
| **uLAM positive (no., %)** | 4/32 (12.5) [4.1 – 29.9] | 3/80 (3.8) [1.0 – 11.3] | 4/51 (7.8) [2.5 – 19.7] | 3/61 (4.9) [1.3 – 14.6] | 1/22 (4.5) [0.2 – 24.9] | 6/90 (6.7) [2.7 – 14.5] |
| **uLAM by HIV status** | | | | | | |
| HIV-positive (no., %) | 1/18 (5.6) [0.3 – 29.4] | 3/45 (6.7) [1.7 – 19.3] | 1/30 (3.3) [0.2 – 19.1] | 3/33 (9.1) [2.4 – 25.5] | 1/13 (7.7) [0.4 – 37.9] | 3/50 (6.0) [1.6 – 17.5] |
| HIV-negative (no., %) | 3/14 (21.4) [5.7 – 51.2] | 0/35 (0.0) [0.0 – 12.3] | 3/21 (14.3) [3.8 – 37.4] | 0/28 (0.0) [0.0 – 15.0] | 0/9 (0.00) [0.0 – 37.1] | 3/40 (7.5) [2.0 – 21.5] |
| **p-value*** | 0.30 | 0.25 | 0.29 | 0.24 | 1.00 | 1.00 |
| **uLAM by CD4⁺ T-cell count[a]** | | | | | | |
| < 200 (no., %) | 1/8 (12.5) [0.0 – 53.3] | 0/12 (0.0) [0.0 – 30.1] | 1/13 (7.7) [0.4 – 37.9] | 0/7 (0.0) [0.0 – 43.9] | 0/8 (0.00) [0.0 – 40.2] | 1/12 (8.3) [0.4 – 40.2] |
| ≥ 200 (no., %) | 0/10 (0.0) [0.0 – 34.5] | 3/33 (9.1) [2.4 – 25.5] | 0/17 (0.0) [0.0 – 22.9] | 3/26 (11.5) [3.0 – 31.3] | 1/5 (20.0) [1.1 – 70.1] | 2/38 (5.3) [0.9 – 19.1] |
| **p-value*** | 0.44 | 0.55 | 0.43 | 1.00 | 0.38 | 1.00 |
| **HRS positive (no., %)** | 15/32 (48.9) [30.9 – 63.6] | 35/80 (43.8) [33.4 – 54.7] | 25/51 (49.0) [35.9 – 62.3] | 25/61 (41.0) [29.5 – 53.5] | 22/22 (100.0) [85.1 – 1.00] | 28/90 (31.1) [22.5 – 41.3] |
| **HRS by HIV status** | | | | | | |
| HIV-positive (no., %) | 11/18 (61.1) [38.6 – 79.7] | 23/45 (51.1) [37.0 – 65.0] | 18/30 (60.0) [42.3 – 75.4] | 16/33 (48.5) [32.5 – 64.8] | 13/13 (100.0) [77.2 – 1.00] | 21/50 (42.0) [29.4 – 55.8] |
| HIV-negative (no., %) | 4/14 (28.6) [11.7 – 54.6] | 12/35 (34.3) [20.8 – 50.8] | 7/21 (33.3) [17.2 – 54.6] | 9/28 (32.1) [17.9 – 50.7] | 9/9 (100.0) [70.1 – 1.00] | 7/40 (17.5) [8.7 – 31.9] |
| **p-value** | 0.09 | 0.17 | 0.09 | 0.30 | 1.00 | **0.02** |
| **HRS by CD4⁺ T-cell count[a]** | | | | | | |

| | | | | | | |
|---|---|---|---|---|---|---|
| < 200 (no., %) | 7/8 (87.5)<br>[52.9 – 97.8] | 10/12 (83.3)<br>[55.2 – 95.3] | 11/13 (84.6)<br>[57.8 – 95.7] | 6/7 (85.7)<br>[48.7 – 97.4] | 8/8 (100.0)<br>[67.6 – 1.00] | 9/12 (75.0)<br>[46.8 – 91.1] |
| ≥ 200 (no., %) | 4/10 (40.0)<br>[16.8 – 68.7] | 13/33 (39.4)<br>[24.7 – 56.3] | 7/17 (41.2)<br>[21.6 – 64.0] | 10/26 (38.5)<br>[22.4 – 57.5] | 5/5 (100.0)<br>[56.6 – 1.00] | 12/38 (31.6)<br>[19.1 – 47.5] |
| **p-value*** | 0.07 | **0.02** | **0.03** | **0.04** | 1.00 | **0.02** |

MRS: microbiological reference criterion. CRS: clinical reference criterion. LCA: latent class analysis. LCM: latent class model.

*Comparison by Fisher's exact test.

[a]Among HIV-positive participants only.

**Table S4.** Selected prior distributions for latent class model parameters and supporting evidence.

| Parameter | Selected Prior Distribution | Interpretation | Evidence/Calculations | Source |
|---|---|---|---|---|
| Ultra sensitivity | Beta(13.90, 16.32) | 95% probability that true value is 0.29 – 0.64. | **0.29** (212/720) of $Sm^-$ and $Cx^-$ patients with a clinical diagnosis of tuberculosis | Feng, 2025[1] |
| | | | **0.29** (22/76) of symptomatic $Cx^-$ children | Peng, 2022[2] |
| | | | **0.64** (7/11) of clinically diagnosed, $Cx^-$ individuals undergoing bronchoscopy on suspicion of PTB | Chien, 2020[3] |
| | | | **0.60** (105/175) of paucibacillary, $Cx^-Sm^-$ specimens from TB suspects | Wang, 2019[4] |
| Ultra specificity | Beta(3.41, 0.59) | The expected value of the specificity is 0.85 (mean of estimates to allow influence of outlier), with a greater probability of higher specificity. | **0.97** (199/206) in paucibacillary, $Sm^-$ sputum from TB suspects | Wang, 2019[4] |
| | | | **0.94** (95% CI = 91.3 – 95.9) pooled estimate among people with $Cx^+Sm^-$ sputum | Horne, 2025[5] |
| | | | **0.97** in $Cx^-$ specimens (144/149) in a low prevalence population | Opota, 2019[6] |
| | | | **0.53** of participants with trace-positive sputum did not progress within 2 years (minimum estimate). 53/168 of study cohort were sputum trace-positive. | Sung, 2026[7] |
| mRNA sensitivity | Beta(1.85, 3.15) | The expected value of the sensitivity is 0.35 (median of the estimates), with a greater probability of low sensitivity.[8] | **0.14** (50/307) TB-presumptive (clinical) $Sm^-$ and $Cx^-$ individuals | Ahmadi Ghezeldasht, 2025[9] |
| | | | Sensitivities greater than **0.60** in smear-negative sputum, higher in smear-positive | Wei, 2019[10] |
| | | | mRNA detected in **35%** of $Cx^-$ sputum after 6 months, 100% of BAL samples | Malherbe, 2016[11] |
| | | | 85B mRNA RT-qPCR assay showed **83%** sensitivity among smear-negative spiked sputum | Jou, 1997[12] |
| | | | **0% - 13%** sensitivity in $Cx^-$ sputum from cohort undergoing TB treatment | Zainabadi, 2022[13] |
| mRNA specificity | Beta(7.12, 1.33) | 95% probability that true value is 0.55 – 0.99. | **Meta-analysis: 92%** pooled specificity, range **0.55 – 1.00** | Wei, 2019[10] |

| | | | | |
|---|---|---|---|---|
| | | | 85B mRNA RT-qPCR assay showed **95.0%** specificity (57/60) for active TB | Demirci, 2018[14] |
| | | | **1.00** (17/17) of RT-PCR positive patients were confirmed by culture and/or improved with treatment | Vinuesa, 2018[15] |
| Longitudinal culture sensitivity | Beta(8.94, 42.01) | 95% probability that true value is 0.10 – 0.29. | **0.23** (19/83) of untreated, radiologically-diagnosed TB patients with all negative initial cultures | Hong Kong Chest, 1981[16] |
| | | | **Meta-analysis: 0.10** (95% CI = 0.06-0.14) from 18 cohorts with abnormal chest X-rays became $Cx^+$ during follow-up | Sossen, 2023[17] |
| | | | **0.29** (88/152 over 2 years) individuals with apical lung legions on CXR and positive tuberculin tests became smear/culture/histologically positive | Cowie, 1985[18] |
| | | | **0.09** per year (46/268 over 2 years) individuals with TB-suggestive CXR became smear positive $Sm^+$ or $Cx^+$ | Okada, 2012[19] |
| uLAM sensitivity | Beta(1,1) | All values are equally likely. (95% probability that true value is 0.025 – 0.975.) | N/A (Uninformative prior) | N/A |
| uLAM specificity | Beta(1,1) | All values are equally likely. (95% probability that true value is 0.025 – 0.975.) | N/A (Uninformative prior) | N/A |
| HRS sensitivity | Beta(1,1) | All values are equally likely. (95% probability that true value is 0.025 – 0.975.) | N/A (Uninformative prior) | N/A |
| HRS specificity | Beta(1,1) | All values are equally likely. (95% probability that true value is 0.025 – 0.975.) | N/A (Uninformative prior) | N/A |
| Prevalence | Beta(2,3) | Prevalence is unlikely to be close to 1.0 (weakly informative prior). | **17%** (152/870) of PTB cases were culture negative | Chen, 2023[20] |
| | | | **15%** (116/796) of PtB cases (clinical & radiographic presentation, improved with treatment) were culture negative | Nguyen, 2019[21] |

| | | | | |
|---|---|---|---|---|
| Random effect | $N_{tr}(0, 1, 0, \infty)$ | The random effect is normally distributed and positive (truncated at 0) with a mean of 0 and a standard deviation of 1. | Random effect of bacterial burden expected to be positive for all tests. | N/A |
| mRNA sensitivity (sensitivity analysis)[b] | Beta(0.77, 4.23) | Median sensitivity of 0.10 | Selected for sensitivity analysis: lower range of estimates | N/A |
| | Beta(2.93, 2.07) | Median sensitivity of 0.60 | Selected for sensitivity analysis: optimistic estimate | N/A |
| | Beta(4.02, 0.98) | Median sensitivity of 0.85 | Selected for sensitivity analysis: upper range of estimates | N/A |

[a]Priors representing the range of possible Ultra specificities include a non-informative prior and two additional priors selected by decreasing the mean of the Ultra specificity estimates from the literature in increments of 0.25.

[b]Priors representing the range of possible mRNA sensitivities were selected by increasing or decreasing the median of the mRNA sensitivity estimates from the literature in increments of 0.25.

**Table S5.** Summary of missing data for indicator tests in the culture-negative cohort (n = 112).

| | Extended MGIT, n (%) | Supplemented MGIT, n (%) | mRNA Transcripts, n (%) | Xpert Ultra at Follow-up, n (%) | Host Response Signature, n (%) | uLAM Assay, n (%) |
|---|---|---|---|---|---|---|
| Results available | 101 (90.2) | 68 (60.7) | 112 (100.0) | 104 (92.9) | 112 (100.0) | 112 (100.0) |
| Results unavailable | 11 (9.8) | 44 (39.3) | 0 (0.0) | 8 (7.1) | 0 (0.0) | (0.0) |
| Treatment initiated before/by month 2 | 0 (0.0) | 26 (23.2) | — | 0 (0.0) | — | — |
| Month-2 follow-up not completed or completed by phone | 0 (0.0) | 3 (2.7) | — | 4 (3.6) | — | — |
| Follow-up completed; result not documented | 0 (0.0) | 11 (9.8) | — | 3 (2.7) | — | — |
| Missingness reason not yet established | 11 (9.8) | 4 (3.6) | — | 1 (0.9) | — | — |

**Table S6.** Concordance between the clinical criterion, the microbiological criterion and the LCM class assignments.

| | Clinical criterion | Microbiological criterion | LCM classification |
|---|---|---|---|
| Clinical criterion | | | |
| Microbiological criterion | 93/112 (0.83) | | |
| LCM classification | 61/112 (0.54) | 72/112 (0.64) | |

**Table S7.** Odds ratios from an exploratory descriptive logistic regression model of all available cases in the training set (n = 128)[a], adjusted for prior history of TB, HIV status, symptom duration and symptom count; and sensitivity, specificity and overall accuracy for model predictions of the test set (n = 56).

| | Microbiological criterion | | Clinical criterion | |
|---|---|---|---|---|
| **Training set** | **OR (95% CI)** | **p-value** | **OR (95% CI)** | **p-value** |
| uLAM result | **5.38 (1.63 – 13.02)** | **0.002**** | 2.36 (0.997 – 5.59) | 0.051 |
| HRS score[b] | 1.04 (0.84 – 1.28) | 0.26 | 1.17 (0.95 – 1.44) | 0.13 |
| HIV positive | 1.04 (0.45 – 2.40) | 0.89 | 1.32 (0.58 – 2.99) | 0.51 |
| Prior TB history | 1.01 (0.38 – 2.66) | 0.39 | 0.91 (0.35 – 2.40) | 0.85 |
| Symptom duration (days)[c] | 1.00 (0.85 – 1.17) | 0.74 | 0.92 (0.76 – 1.10) | 0.35 |
| Total number of symptoms | **1.74 (1.16 – 2.60)** | **0.002**** | 1.36 (0.93 – 1.99) | 0.11 |

**Test set**

| Criterion | OR [95% CI] | Fisher Test p-value | Sensitivity [95% CI] | Specificity [95% CI] | Accuracy [95% CI] |
|---|---|---|---|---|---|
| Microbiological | **3.74 (1.07 – 14.26)** | **0.03*** | 0.54 (14/26) [0.35 – 0.71] | 0.77 (23/30) [0.59 – 0.88] | 0.66 (37/56) [0.53 – 0.77] |
| Clinical | 1.49 (0.32 – 7.16) | 0.74 | 0.83 (25/30) [0.66 – 0.93] | 0.23 (6/26) [0.11 – 0.42] | 0.55 (31/56) [0.42 – 0.68] |

*p < 0.05, **p < 0.01, ***p < 0.001

[a]Culture-positive participants are included as reference positive and ORC participants are included as reference negative.

[b]Composite score constructed from standardized ACS-COR and standardized and reverse-coded PREDICT29 HRS scores.

[c]Defined as the reported duration of coughing in Ultra participants, and the reported time between first symptoms and screening in ORC participants. TB symptoms include weight loss, night sweats and hemoptysis.

**Table S8.** Sensitivity and specificity estimates of all indicator tests for four latent class models.

| | **Model 1: Base Model** | | **Model 2: uLAM Index** | | **Model 3: HRS Index** | | **Model 4: Full Model** | |
|---|---|---|---|---|---|---|---|---|
| Est. prevalence (95% CrI) | 0.37 (0.10 – 0.75) | | 0.35 (0.08 – 0.76) | | 0.31 (0.12 – 0.60) | | 0.31 (0.11 – 0.64) | |
| Positive classification (%) | 6/112 (5) | | 7/112 (6) | | 42/112 (38) | | 22/112 (20) | |
| | **Sensitivity** | **Specificity** | **Sensitivity** | **Specificity** | **Sensitivity** | **Specificity** | **Sensitivity** | **Specificity** |
| Xpert Ultra | 0.47 (0.31 – 0.64) | 0.69 (0.52 – 0.98) | 0.46 (0.31 – 0.62) | 0.67 (0.52 – 0.96) | 0.45 (0.31 – 0.60) | 0.65 (0.52 – 0.80) | 0.45 (0.30 – 0.60) | 0.65 (0.51 – 0.81) |
| mRNA Transcripts | 0.21 (0.03 – 0.50) | 0.78 (0.61 – 0.95) | 0.24 (0.04 – 0.62) | 0.79 (0.58 – 0.95) | 0.13 (0.02 – 0.34) | 0.74 (0.60 – 0.85) | 0.13 (0.02 – 0.42) | 0.74 (0.58 – 0.87) |
| Longitudinal culture[a] | 0.15 (0.08 – 0.26) | 0.98 | 0.15 (0.08 – 0.26) | 0.98 | 0.17 (0.09 – 0.28) | 0.98 | 0.17 (0.09 – 0.28) | 0.98 |
| uLAM Immunoassay | — | — | 0.09 (0.00 – 0.48) | 0.94 (0.79 – 1.00) | — | — | 0.06 (0.00 – 0.28) | 0.92 (0.82 – 0.98) |
| HRS assay | — | — | — | — | 0.82 (0.42 – 0.99) | 0.71 (0.51 – 0.95) | 0.78 (0.34 – 0.99) | 0.69 (0.48 – 0.93) |

[a]Longitudinal culture specificity set at 0.98.

**Table S9.** Prevalence estimation, class assignment and sensitivity and specificity estimates of index tests under different assumed priors for mRNA sensitivity.

| **mRNA sensitivity** | **Prior 1: beta(0.77,4.23)** | | **Prior 2: beta(1.85,3.15)** | | **Prior 3: beta(2.93,2.07)** | | **Prior 4: beta(4.02,0.98)** | |
|---|---|---|---|---|---|---|---|---|
| Est. prevalence (95% CrI) | 0.30 (0.12 – 0.58) | | 0.31 (0.11 – 0.64) | | 0.29 (0.06 – 0.65) | | 0.16 (0.03 – 0.56) | |
| Positive classification (%) | 35/112 (31) | | 22/112 (20) | | 7/112 (6) | | 4/112 (4) | |
| | **Sensitivity** | **Specificity** | **Sensitivity** | **Specificity** | **Sensitivity** | **Specificity** | **Sensitivity** | **Specificity** |
| mRNA Transcripts | 0.05 (0.00 – 0.24) | 0.72 (0.56 – 0.82) | 0.13 (0.02 – 0.42) | 0.74 (0.58 – 0.87) | 0.22 (0.06 – 0.76) | 0.77 (0.61 – 0.92) | 0.63 (0.13 – 0.99) | 0.82 (0.68 – 0.95) |
| uLAM Immunoassay | 0.05 (0.00 – 0.21) | 0.92 (0.82 – 0.97) | 0.06 (0.00 – 0.28) | 0.92 (0.82 – 0.98) | 0.08 (0.00 – 0.54) | 0.93 (0.83 – 0.99) | 0.21 (0.01 – 0.88) | 0.95 (0.86 – 0.99) |
| HRS assay | 0.77 (0.43 – 0.99) | 0.68 (0.51 – 0.88) | 0.78 (0.34 – 0.99) | 0.69 (0.48 – 0.93) | 0.72 (0.16 – 0.99) | 0.66 (0.44 – 0.93) | 0.43 (0.03 – 0.97) | 0.56 (0.42 – 0.85) |

**Figure S1.** ROC-AUC curves for the (a) S4-20 IgM, (b) PREDICT29 HRS and (c) ACS-COR HRS biomarkers used to set independently derived cutoffs.

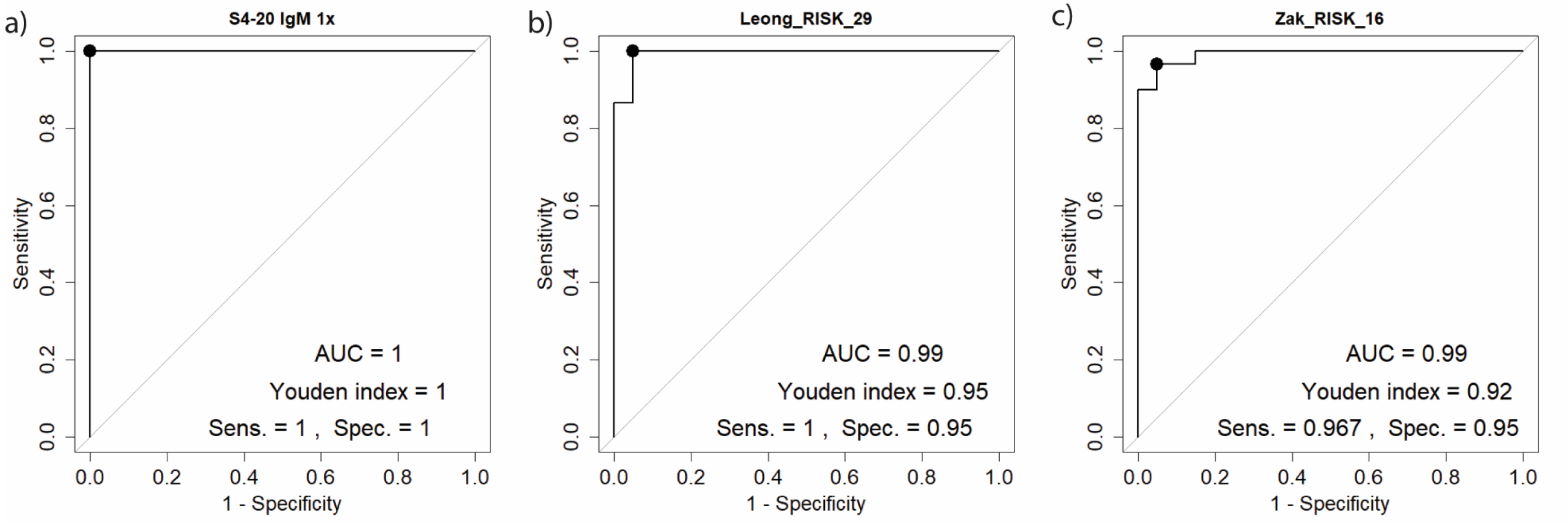

Alt text: Graphical representation of ROC-AUC curves used in setting independently derived cutoffs for biomarkers, with subfigures (a) for S4-20 IgM, (b) for PREDICT29 HRS and (c) for ACS-COR HRS. Figures display high biomarker sensitivity and specificity with AUCs at or approaching 1.0.

**Figure S2.** ROC-AUC curves for the (a) S4-20 IgM, (b) PREDICT29 HRS and (c) ACS-COR HRS biomarkers based on the culture positive samples as positives and ORC and healthy volunteers as negatives.

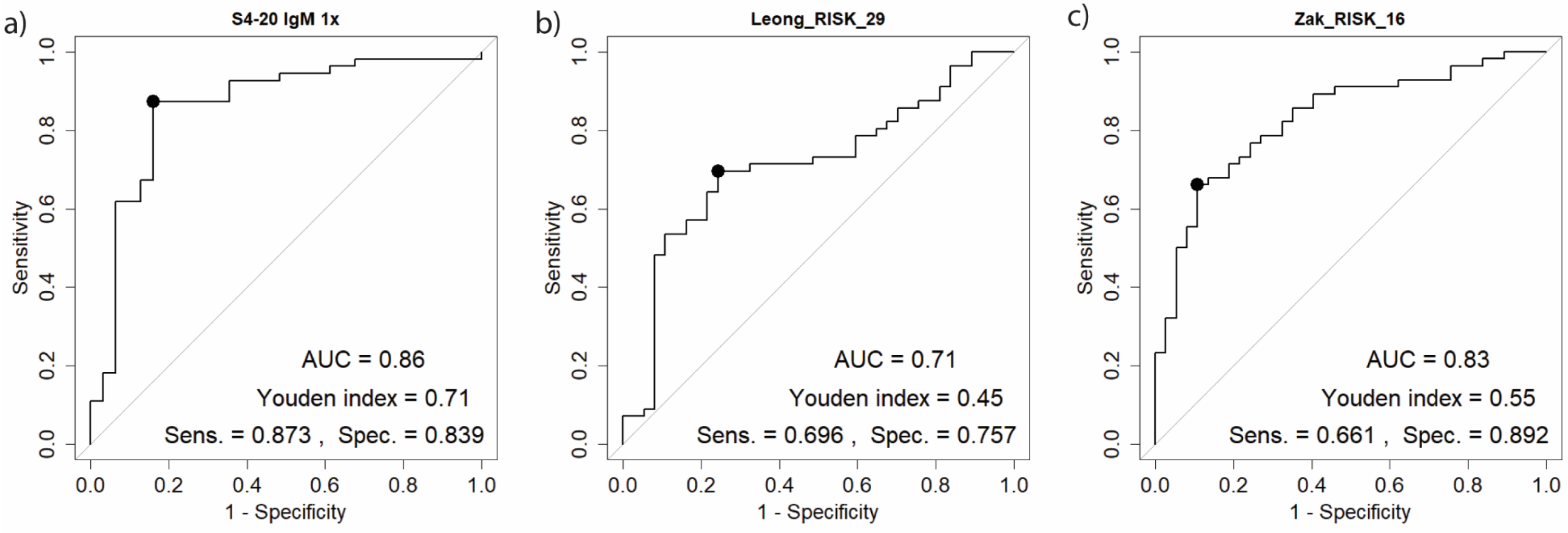

Alt text: Graphical representation of recalculated ROC-AUC curves for separating $Cx^+$ participants from ORC and healthy volunteers, with subfigures (a) for S4-20 IgM, (b) for PREDICT29 HRS and (c) for ACS-COR HRS. Figures display moderate biomarker sensitivity, with and specificity with AUCs of 0.86 for S4-20 IgM, 0.71 for PREDICT29 HRS and 0.83 for ACS-COR HRS.

**Figure S3.** Directed acyclic graph illustrating the relationships between indicator assays (including index tests) and the hypothesized latent classes.

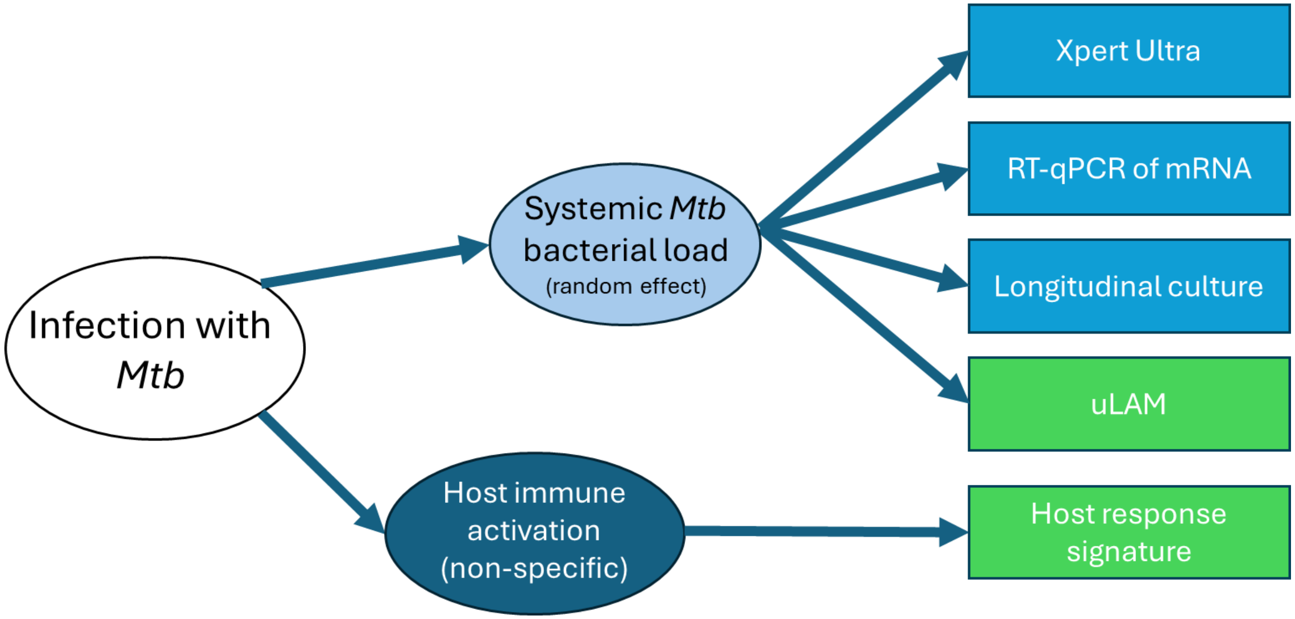

Alt text: A directed graph of the relationship between latent classes and indicator assays. Mtb infection status gives rise to systemic bacterial load, which determines the results of the Ultra, mRNA, longitudinal culture and uLAM assays, and to host immune activation, which determines the host response signature.

**Figure S4.** Venn diagram of positive uLAM and HRS biomarkers, by cohort, for (a) culture-positive cohort (including $Cx^{+}Sm^{+}$ and $Cx^{+}Sm^{-}$), (b) culture-negative cohort $Cx^{-}$ in total, (c) other respiratory diseases cohort ORC, (d) culture-negative cohort $Cx^{-}$ by the microbiological reference standard, (e) culture-negative cohort $Cx^{-}$ by the clinical reference standard, (f) cohort $Cx^{-}$ by the LCA class assignment.

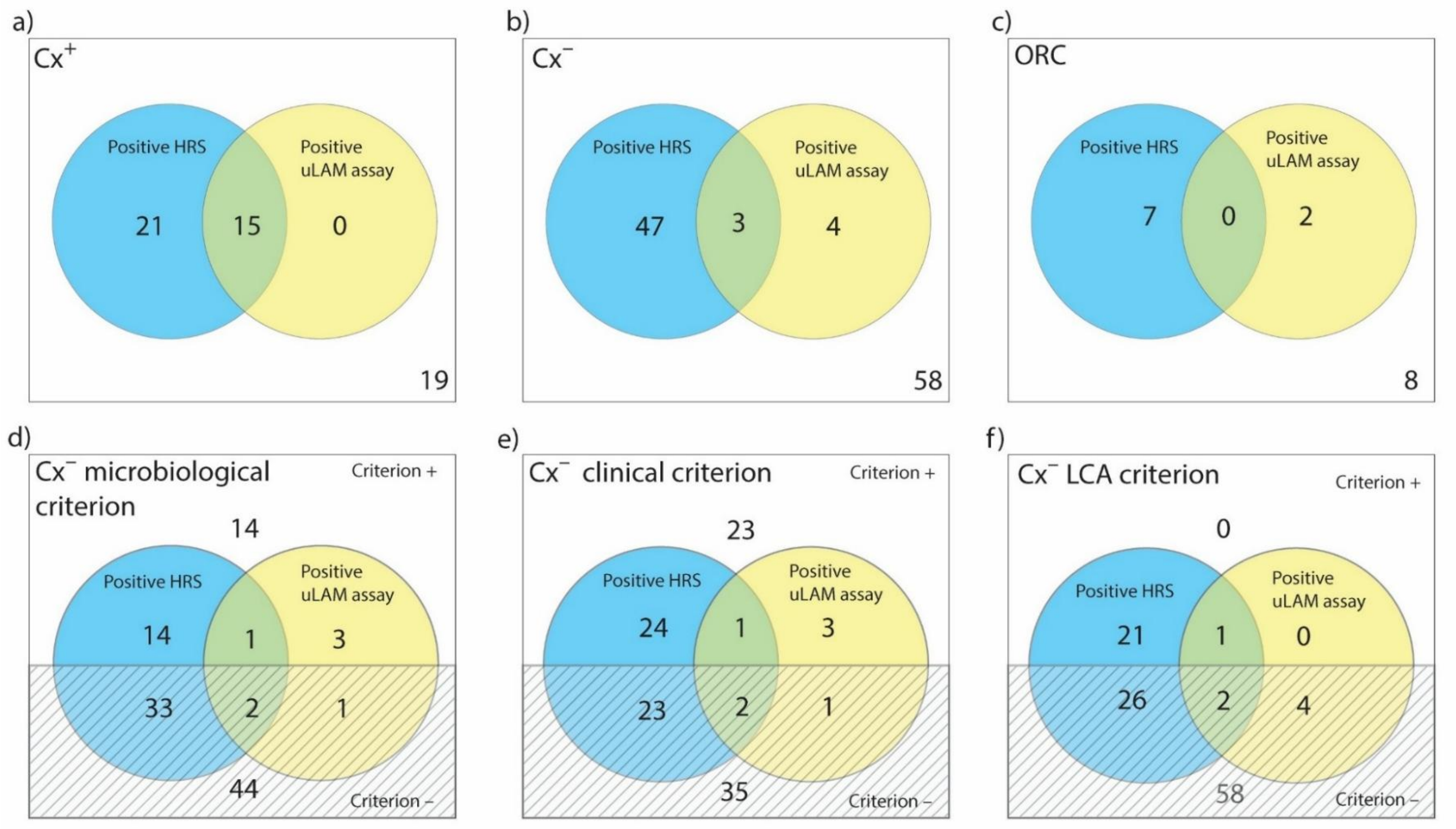

Alt text: Venn diagrams of uLAM and HRS assay results in six panels, for (a) the total $Cx^{+}$ cohort, (b) the $Cx^{-}$ cohort, (c) the ORC cohort, (d) the the $Cx^{-}$ cohort by MRS classification, (e) the $Cx^{-}$ cohort by CRS classification and (f) the $Cx^{-}$ cohort by LCA classification.

**Figure S5.** (a) Venn diagram of overlapping biomarkers and (b) upset plot of biomarker patterns in the Cx⁻ cohort comparing participant classification by the CRC and the LCM (n = 101)[a].

(a)

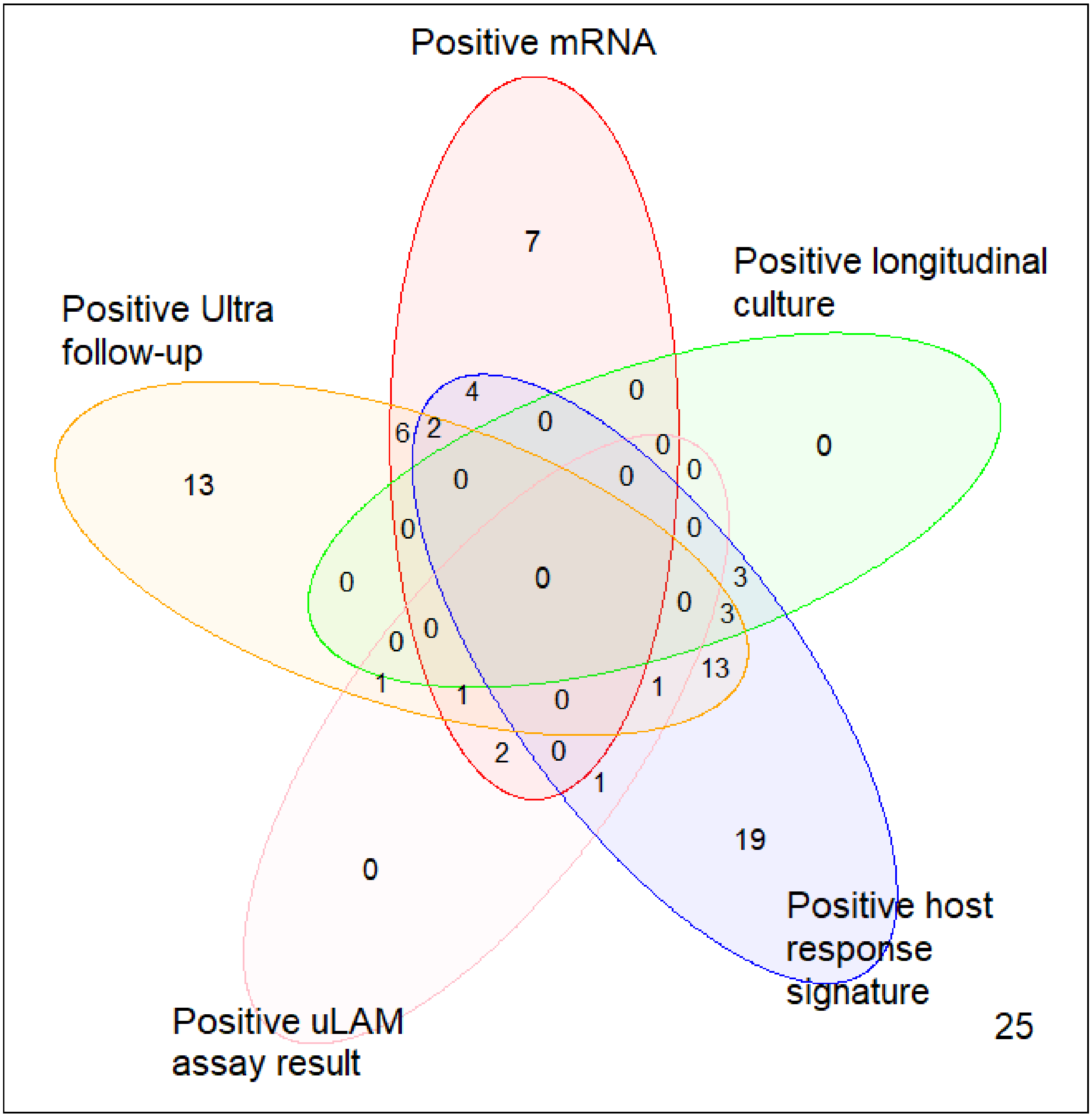

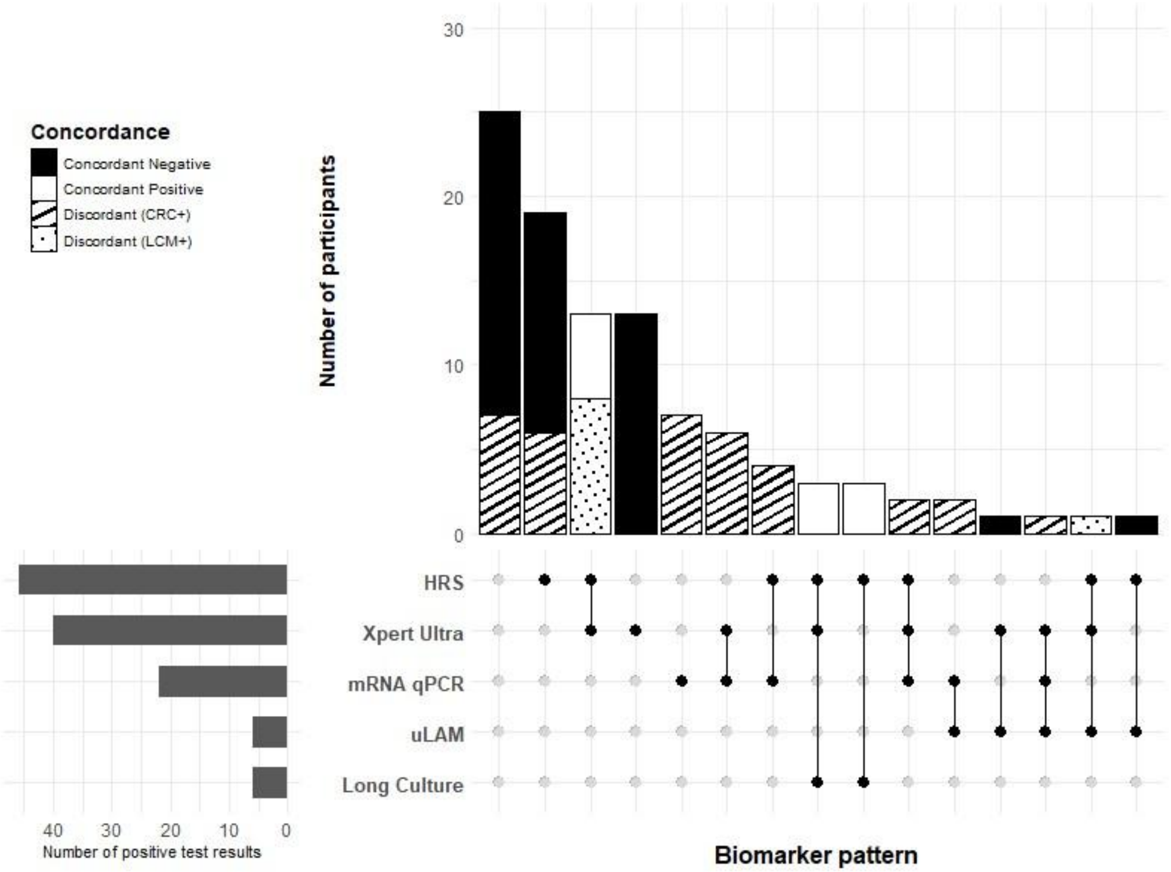

(b)

[a]Excluding 4 participants missing follow-up Ultra results only, 3 missing longitudinal culture results only, and 4 missing both.

Alt Text: A two-panel graphical representation of unique biomarker patterns. Panel (a) shows a five-lobed Venn diagram of biomarker positivity in the $Cx^{-}$ cohort for longitudinal culture, HRS, uLAM, follow-up Ultra and mRNA assays. Overlapping regions indicate unique biomarker patterns. Panel (b) shows a

**Figure S6.** Violin plots of other measured host response signature scores, by cohort.

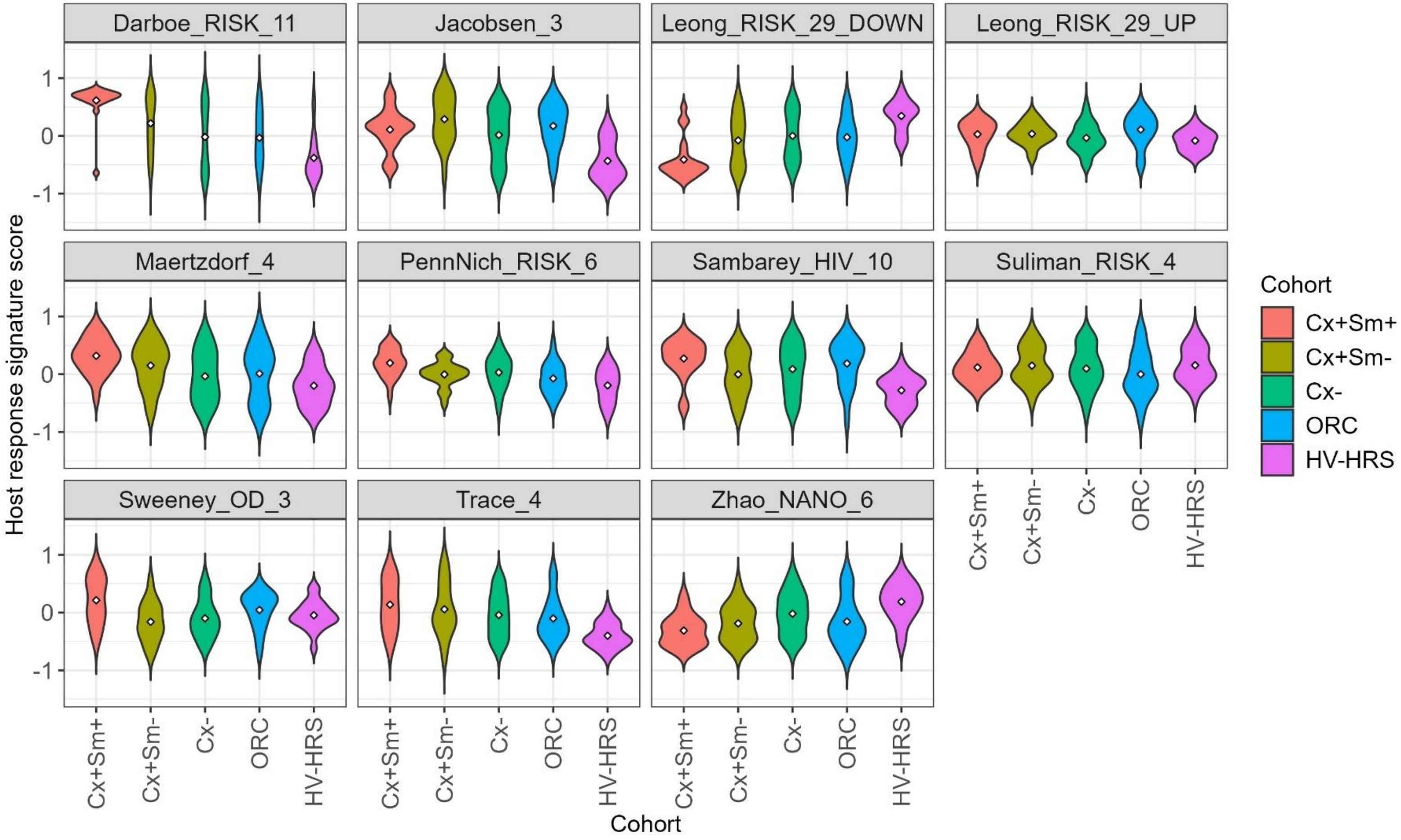

$Cx^-$: sputum culture negative. $Cx^+Sm^+$: sputum culture positive, smear positive. $Cx^+Sm^-$: sputum culture positive, smear negative. ORC: other respiratory condition. HV-HRS: healthy volunteers.

Alt Text: Violin plots showing the host response signature scores for the different cohorts across 11 host response signatures that were analyzed but not used in reference criteria or latent class modeling.

**Figure S7.** Violin plots of host response signature scores for the PREDICT29 and ACS-COR signatures, by cohort.

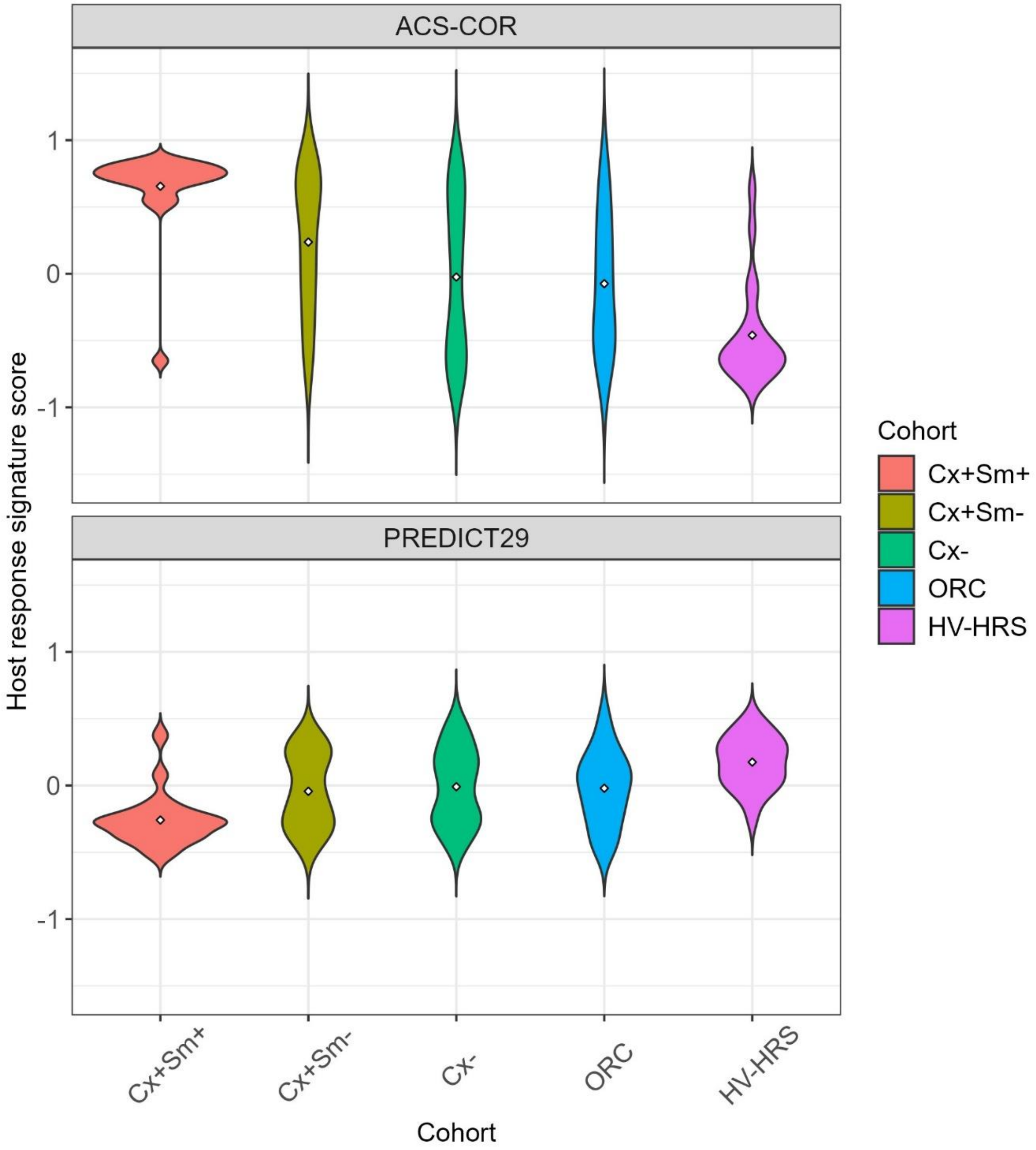

Cx$^-$: sputum culture negative. Cx$^+$Sm$^+$: sputum culture positive, smear positive. Cx$^+$Sm$^-$: sputum culture positive, smear negative. ORC: other respiratory condition. HV-HRS: healthy volunteers.

Alt Text: Violin plots showing the host response signature (HRS) scores for the different cohorts across the ACS-COR and PREDICT29 host response signatures used in reference criteria and latent class modeling to determine positivity. ACS-COR scores decrease and PREDICT29 scores increase with decreasing assumed bacterial burden.

**Figure S8.** Boxplots of uLAM immunoassay signal-to-noise ratios, by cohort.

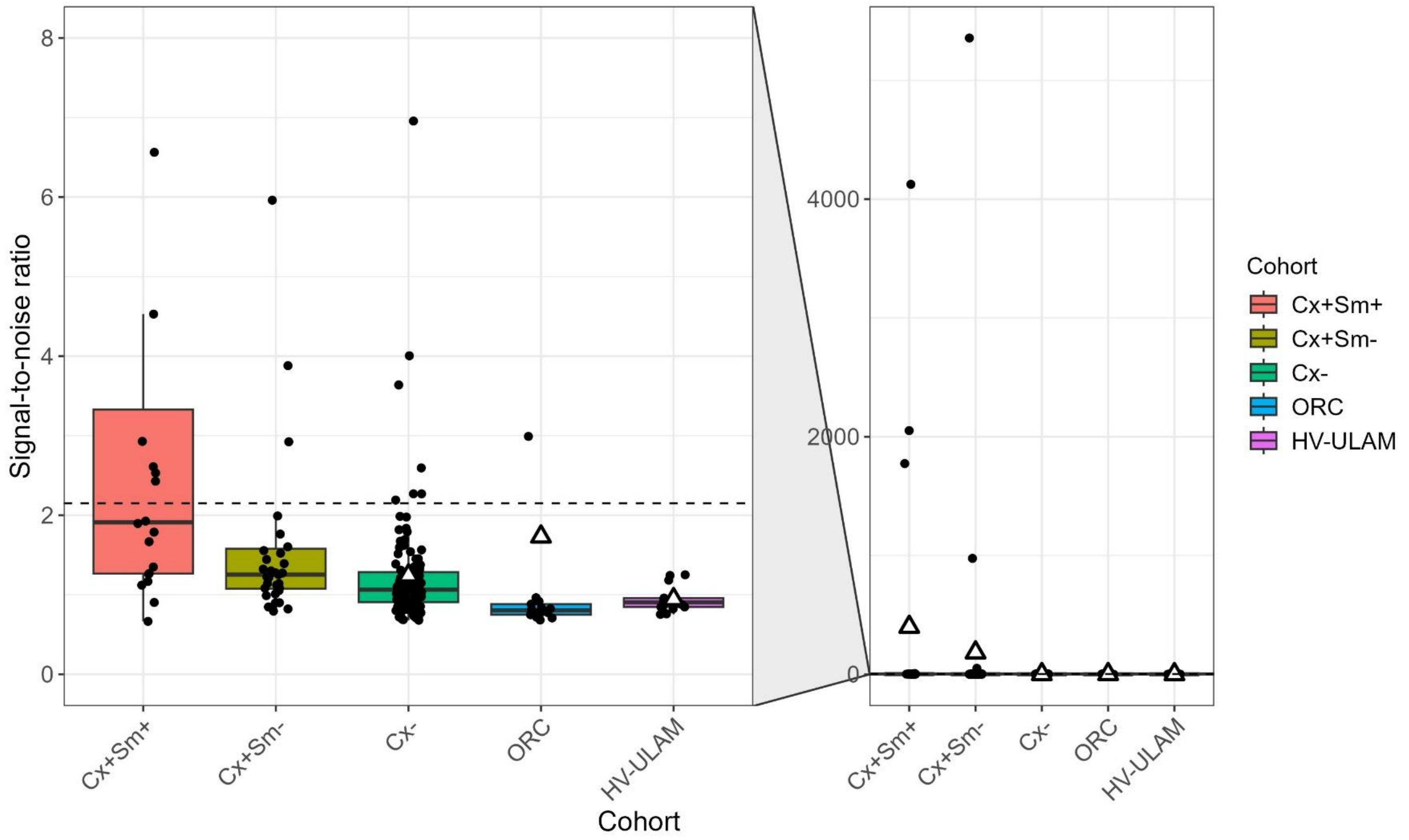

Cx$^{-}$: sputum culture negative. Cx$^{+}$Sm$^{+}$: sputum culture positive, smear positive. Cx$^{+}$Sm$^{-}$: sputum culture positive, smear negative. ORC: other respiratory condition. HV-ULAM: healthy volunteers.

Alt text: Graphical representation of uLAM assay signal-to-noise ratio by cohort, with a panel showing outliers. Signal-to-noise ratio generally decreases with decreasing assumed bacterial burden, being higher in culture-positive cohorts and lowest in the other respiratory conditions and healthy volunteers.

**Figure S9.** Kernel density plots of Markov Chain Monte Carlo draws for parameters of interest.

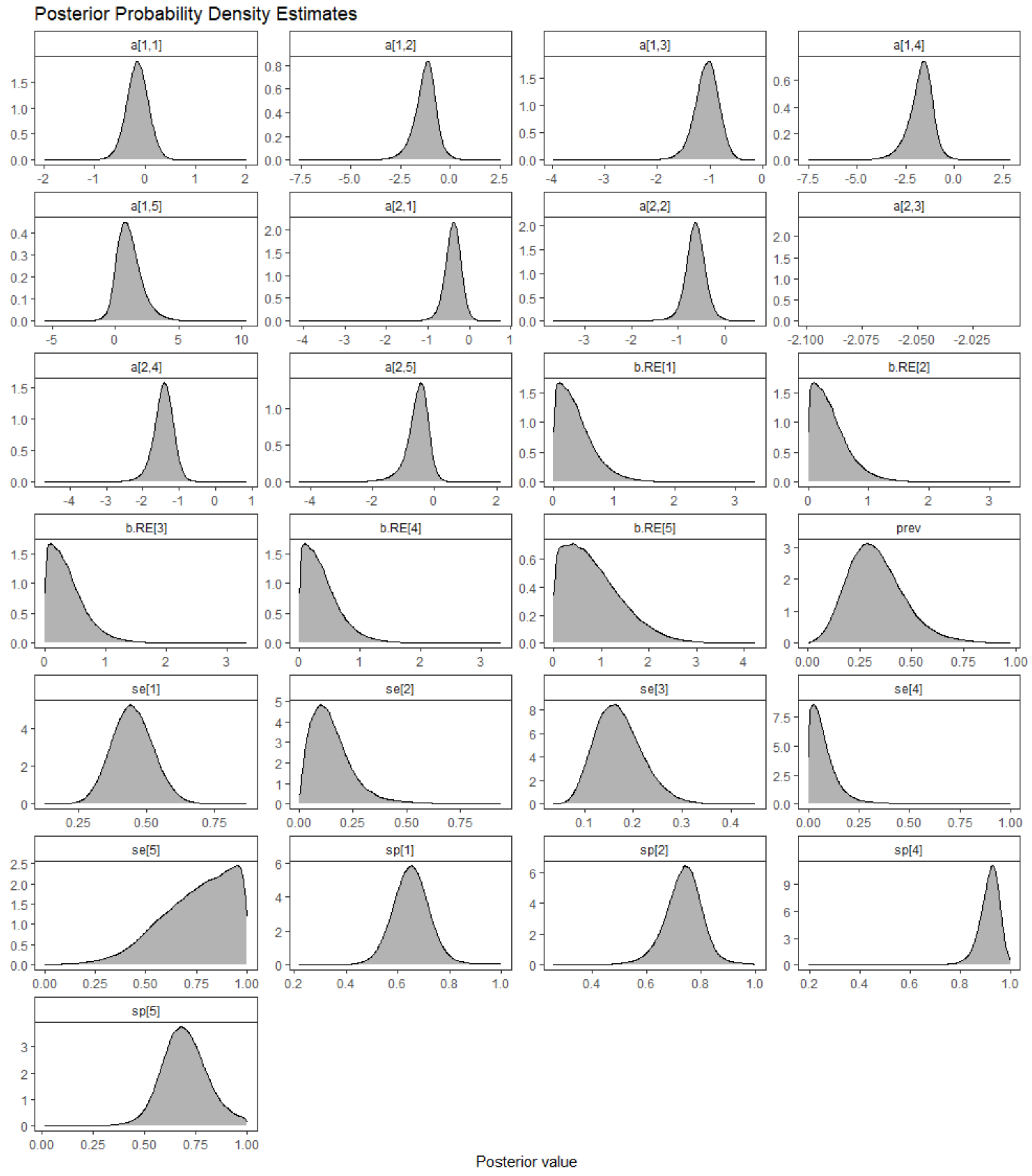

Alt text: Panels displaying kernel density plots for LCM posterior distribution draws for all parameters. Most panels display bell-shaped curves except for a[2,3] which is blank and se[5] which shows a left-skewed distribution.

**Figure S10.** Expanded study flowchart (STARD) of selection of participants for the cohort of interest, positive controls, ORC negative controls and samples used for cutoff derivation.

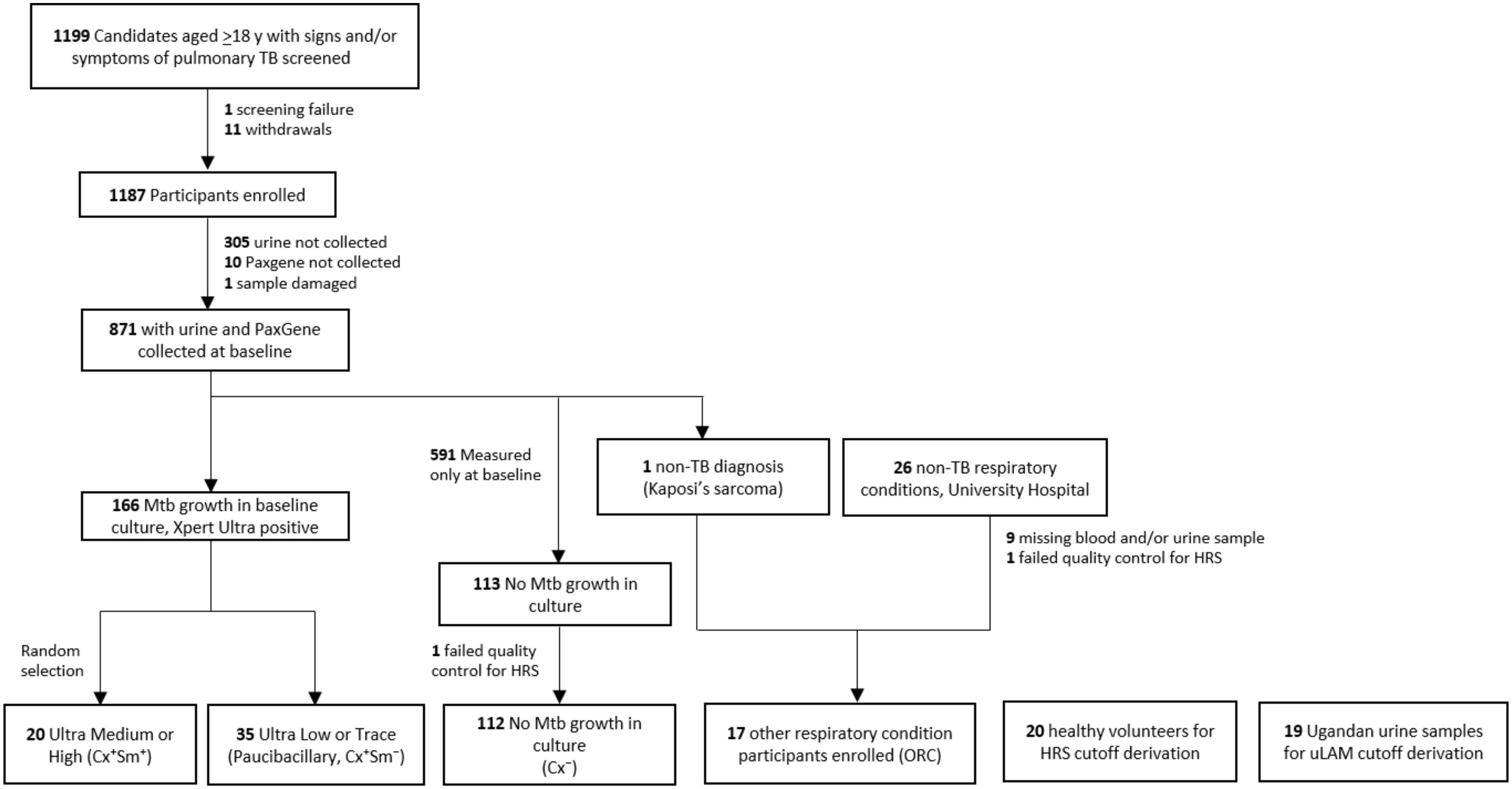

Alt text: An expanded STARD flowchart depicting study participants who were screened (n = 1199), enrolled (n = 1187), had samples collected (n = 871) and were assigned to $Cx^+Sm^+$ (n = 20), $Cx^+Sm^-$ (n = 35), $Cx^-$ (n = 112) or ORC (n = 17) cohorts. The chart also includes healthy volunteer samples used to derive the HRS cutoff (n = 20) and the independent urine samples used to derive the uLAM cutoffs.
